# Peripheral GFAP Predicts Incident Dementia in Parkinson’s Disease

**DOI:** 10.64898/2026.07.07.26357445

**Authors:** Ludo van Hillegondsberg, Kaushik Renganaath, Tanja Zerenner, Karolien Groenewald, Jamil Razzaque, Ayan Ianniello, Paolo Piazza, Richard Wade-Martins, Avigail Taylor, Alexander Thompson, Yoav Ben-Shlomo, Michele Hu

## Abstract

**Importance:** Dementia is a common and disabling complication of Parkinson disease (PD). Blood-based biomarkers that identify individuals at higher risk of future dementia could improve prognostication and trial stratification.

**Objective:** To determine whether blood-based proteins are associated with future risk of dementia in PD.

**Design, Setting, and Participants:** Prospective longitudinal cohort study with discovery and replication analyses in the Oxford Parkinson’s Disease Centre (OPDC) Discovery cohort and the Parkinson’s Progression Markers Initiative (PPMI). A total of 1,335 participants with PD and 431 healthy controls, with serum, plasma, or cerebrospinal fluid proteomic data and follow-up of up to 12 years, were included.

**Main Outcomes and Measures:** Incident dementia, defined using a composite of MoCA scores, MDS-UPDRS items, and clinician diagnosis. Associations between baseline protein levels and time to dementia were evaluated.

**Results:** Among 1,335 participants with PD, 168 developed incident dementia across cohorts (OPDC Discovery [serum], n = 108; PPMI Project 293 [plasma], n = 23; PPMI Project 181 [CSF], n = 37). In the OPDC cohort, GFAP was the only protein (of 5,408 tested) significantly associated with incident dementia (HR = 2.43; 95% CI: 1.79–3.30; *p-adjust* =1.35 × 10^-4^). Higher GFAP tertiles were associated with greater cumulative dementia incidence. Findings were replicated in the PPMI plasma project (HR = 2.42; 95% CI: 1.12–5.22; *p* = 0.024) but not in the CSF project (HR = 0.96; 95% CI: 0.66–1.39; *p* = 0.82). Higher baseline GFAP was significantly associated with lower baseline cognitive performance and greater longitudinal cognitive decline but no significant association with motor progression. In both cohorts, GFAP levels increased over time but showed no group-level differences.

**Conclusions and Relevance:** Circulating GFAP is a robust and reproducible predictor of future dementia in PD, detectable early in the disease course. While the lack of differential longitudinal trajectories between PD patients with and without dementia suggests that GFAP does not act as a dynamic marker of cognitive decline, its relative stability supports its role as an early indicator of underlying biological vulnerability or subclinical pathology. These findings support serum GFAP as a promising, accessible biomarker for early dementia risk stratification.

**Key Points:** *Question:* Are any blood-based proteins associated with future risk of dementia in Parkinson’s disease?

*Findings:* In this prospective longitudinal cohort study of 1,335 participants with Parkinson’s disease, baseline glial fibrillary acidic protein was the only one of 5,408 proteins associated with incident dementia in the discovery cohort and was replicated in an independent plasma cohort, but not in cerebrospinal fluid.

*Meaning:* Peripheral glial fibrillary acidic protein may help identify people with Parkinson’s disease at higher risk of future dementia early in the disease course.

## Introduction

Parkinson’s disease (PD) is a neurodegenerative disorder characterised by the loss of dopaminergic neurons in the substantia nigra and intraneuronal accumulation of misfolded alpha-synuclein in the brain. It is a progressive, multisystem disease characterized by a broad spectrum of motor and non-motor manifestations affects approximately 150 per 100 000 people worldwide.^1,2^ Among these, dementia represents one of the most common and clinically significant non-motor complications, with risk increasing substantially as the disease advances. Cross-sectional studies suggest that approximately 24 – 31% of people with PD have dementia at any given time, while longitudinal cohort studies demonstrate 30–50% developing dementia within 10 years of diagnosis.^3–5^ The annual incidence of Parkinson’s disease dementia (PDD) is estimated at approximately 4–5%, corresponding to a three-to fourfold increased risk compared with age-matched controls.^4–6^

PDD is clinically characterized by deficits in attention, memory and executive and visuospatial function. This is frequently accompanied by cognitive fluctuations, visual hallucinations, and other neuropsychiatric symptoms such as anxiety, depression and apathy.^7^ Clinically and pathologically, PDD overlaps substantially with dementia with Lewy bodies, although by definition motor symptoms must precede the onset of dementia by at least one year in PDD.^7,8^ While cholinesterase inhibitors and memantine may offer temporary symptomatic benefit in PDD, no disease-modifying treatments exist.^9^

The high cumulative prevalence of dementia in PD has major clinical and societal implications. Dementia is a leading determinant of disability, reduced quality of life, caregiver burden, institutionalization, and mortality among people with PD.^6,10^ Older age, longer disease duration, greater disease severity, APOEε4 status, non-motor symptom burden, low Aβ42/40 ratio, elevated glial fibrillary protein and p-tau have been identified as risk factors or markers of future dementia, although rates of progression can still vary substantially between individuals.^11–23^ This heterogeneity has important implications for prognosis, patient counselling, health-care planning, and the development of preventive and disease-modifying strategies, underscoring the need for reliable biomarkers that can identify individuals at increased risk of future dementia early in the disease course.

## Materials and methods

### Study Population

#### OPDC Discovery Cohort

This study drew on the Discovery Cohort of the Oxford Parkinson’s Disease Centre (OPDC), a community-based longitudinal study comprising approximately 1,900 participants recruited over a 14-year period, predominantly from the Thames Valley region of the UK. The cohort includes three principal groups: individuals with PD, those with idiopathic rapid eye movement sleep behaviour disorder (iRBD), and healthy controls. Participants with PD were diagnosed by an NHS neurology consultant prior to enrolment, with their diagnosis confirmed at enrolment by a neurology consultant or fellow using the UK Brain Bank Criteria in a structured interview and examination.^24^ Participants were reviewed every 12–18 months, and those who developed atypical parkinsonian syndromes were withdrawn.

Healthy controls exhibited no evidence of neurodegenerative disease at baseline neurological review, had no first-degree relatives with PD, underwent routine monitoring every 12–18 months and were withdrawn if a neurodegenerative disease developed. Full details of the clinical protocol have been published elsewhere.^26^ The cohort study received approval from the local research ethics committee, and all participants provided written informed consent.

#### PPMI Cohort

Parkinson’s Progression Markers Initiative (PPMI) is an ongoing, international, observational study conducted across the United States, Europe, Africa, Israel, and Australia. To date, over 5,500 participants have been enrolled, including healthy controls, individuals with de novo PD, prodromal PD participants and non-manifesting carriers of LRRK2 and GBA variants. Participants undergo comprehensive clinical assessments, imaging, and molecular phenotyping every 6-12 months. Study protocol and manuals are available at https://www.ppmi-info.org/study-design. For this study we analysed data from two PPMI sub-studies (Project 293 & Project 277), conducted by industry research groups in collaboration with PPMI and made available through the PPMI data portal under the data use agreement. Data used in this article were obtained on 13 March 2026, from the PPMI database (https://www.ppmi-info.org/access-data-specimens/data), RRID:SCR_006431.

#### Proteomic analysis and quality control

Proteomic analysis was performed using proximity extension assay (PEA) technology (Olink® Proteomics, Uppsala, Sweden). This approach uses pairs of target-specific antibodies, each conjugated to a unique oligonucleotide corresponding to a protein in the panel. Upon binding to their target protein, the paired oligonucleotides hybridize and are subsequently amplified by polymerase chain reaction. The resulting amplicons are quantified using next-generation sequencing and expressed as normalized protein expression (NPX) values on a logL scale, representing relative protein abundance.^27^

The proteomic analysis of the OPDC Discovery cohort was conducted by an independent team at the Centre for Human Genetics, University of Oxford, who were blinded to all clinical data. All serum were analysed with the Olink® Explore HT platform. Sample plate layouts were randomized, with each plate layout programmatically generated using the concept of negative spatial autocorrelation.^28^ Raw data acquisition and quality control (QC) followed the manufacturer’s protocol. ^29,30^ The intensity-normalised NPX values were utilized, and additional QC steps were employed to identify sample outliers and calculate assay performance metrics. Of 5,408 assays that passed QC, 1,810 (33.47%) had <30% of results below the assay-specific lower limit of detection (LOD). This subset of assays demonstrated a mean interplate coefficient of variance (CV) of 11.47%. Nevertheless, in line with our exploratory approach, all 5,408 assays were retained for downstream analysis, as subtle group-level differences may still be detectable and any LOD threshold is inherently somewhat arbitrary. Further details on the QC steps can be found in the supplementary material (**Supplementary Figures 1 & 2, 5, Supplementary Table 1**).

For the PPMI cohort, study documents describing Project 293 and Project 277 are available here: https://ida.loni.usc.edu/pages/access/studyData.jsp?categoryId=7&subCategoryId=305.

Briefly, Project 293 and Project 277 were large-scale Olink® Explore HT proteomics studies. Project 293 tested plasma samples from Parkinson’s disease participants, prodromal individuals, and healthy controls, totalling 874 samples from 304 participants. Project 277 tested CSF samples from the same three arms of the cohort, totalling 1200 samples from 746 participants. Raw data acquisition and quality control (QC) followed the manufacturer’s protocol. ^29,30^ Only the relevant protein’s intensity-normalised NPX values were utilized in this article, and additional QC step was performed to identify sample outliers (**Supplementary Figures 3 & 4**)

## Statistical Analysis

Demographic characteristics of the study population were analysed to assess baseline differences. Distributions of continuous variables across groups were compared using the Kruskal-Wallis test and binary variables were evaluated with Pearson’s Chi-squared test. Time-to-event analyses were conducted in PD participants without dementia at baseline and with at least one follow-up visit using multivariable Cox proportional hazards regression models with delayed entry. A delayed entry (left-truncated) Cox regression approach was used to account for blood sampling at enrolment rather than at PD diagnosis, concurrently also adjusting for heterogeneity in disease duration at study entry. Dementia was defined as: (i) an education-adjusted MoCA score < 21 together with a MDS-UPDRS item 1.1 score ≥ 1 and a MDS-UPDRS item 1.3 < 4 at two consecutive visits; (ii) an education-adjusted MoCA score < 21 together with a MDS-UPDRS item 1.1 score ≥ 1 and a MDS-UPDRS item 1.3 < 4 at a single visit if it was the participant’s final assessment; or (iii) a clinical diagnosis of dementia made by either a study clinician or an external NHS clinician. Participants that met dementia criteria at baseline were excluded. Each of the 5,408 proteins was tested individually for association with incident dementia, adjusting for age, sex, body mass index (BMI), and sample age. In secondary analyses, Kaplan–Meier survival curves were generated to illustrate cumulative dementia incidence across baseline protein abundance tertiles. Cross-sectional associations between protein abundance and clinical and genetic variables (age, sex, BMI, APOE ε4 carrier status, MoCA score and MDS-UPDRS III) were evaluated using linear regression models and adjusted for the relevant covariates. Group differences in baseline protein abundance were assessed using linear regression models adjusted for age, sex, disease duration, BMI, and sample age. Longitudinal changes in protein abundance were examined using linear mixed-effects models, with time from baseline, disease duration, age, sex, BMI at baseline, and sample age as fixed effects, and participant ID specified as a random intercept to account for within-individual repeated measures. Longitudinal associations between protein abundance and cognitive trajectories were assessed using linear mixed-effects models incorporating a time × GFAP interaction term, with baseline age, sex, BMI, APOE ε4 carrier status, disease duration and sample age as fixed effects and a random intercept for participant ID to account for within-subject correlation. Where applicable, the p-value was adjusted using the Benjamini-Hochberg false-discovery rate (FDR) correction method and considered significant at < 0.05. All statistical analyses were performed using R (version 4.5.3). Cox proportional hazard models were fitted using the *survival* package (version 3.7.0) and linear mixed effects models were implemented with the *lme4* package (version 2.0.1).

## Results

### Study demographics

Demographic and clinical characteristics are summarised in **Table 1**. A total of 1,766 individuals were included from the OPDC Discovery and PPMI cohorts (healthy controls = 431; PD = 1,335). Of the PD participants, 261 contributed CSF samples (PPMI Project 277), while the remainder contributed plasma or serum samples. PD participants were slightly older than healthy controls and more frequently male. Across PD cohorts, disease duration at baseline was short (mean ≤1.3 years), representing early-stage disease. MDS-UPDRS III scores were highest in the OPDC cohort compared to PPMI cohorts, while MoCA scores were modestly lower in the OPDC PD cohort relative to PPMI cohorts. Baseline dementia prevalence was low in the OPDC Discovery cohort (5%) and absent in PPMI participants, with similar rates of incident dementia observed during follow-up across cohorts (11–14%). The number of participants meeting each component of the dementia classification criteria across datasets is shown in **Supplementary Figure 6**.

**Table 1.** Study population demographics.

|  | <u>OPDC Discovery Cohort</u><br>(Serum) |  | <u>PPMI Cohort (P-293)</u><br>(Plasma) |  | <u>PPMI Cohort (P-277)</u><br>(CSF) |  | p-value |
| --- | --- | --- | --- | --- | --- | --- | --- |
|  | HC<br>(n = 212) | PD<br>(n = 893) | HC<br>(n = 88) | PD<br>(n = 181) | HC<br>(n = 131) | PD<br>(n = 261) |  |
| Age (years) | 65 ± 6 | 67 ± 10 | 60 ± 4 | 60 ± 9 | 61 ± 11 | 62 ± 9 | <0.001 |
| Sex (no. male, %) | 109 (51%) | 568 (64%) | 51 (58%) | 114 (63%) | 84 (64%) | 177 (67%) | 0.002 |
| Years from diagnosis to baseline | - | 1.3 ± 1 | - | 0.8 ± 1 | - | 0.5 ± 0.6 | <0.001 |
| Baseline MDS-UPDRS III | 1 ± 2 | 27 ± 11 | 1 ± 2 | 19 ± 8 | 1 ± 2 | 20 ± 8 | <0.001 |
| Hoehn & Yahr stage (no., %) |  |  |  |  |  |  |  |
| 1 | - | 204 (23%) | - | 96 (53%) | - | 117 (45%) |  |
| 2 | - | 614 (69%) | - | 84 (46%) | - | 144 (55%) |  |
| 3 | - | 69 (7.8%) | - | - | - | - |  |
| 4-5 | - | 1 (0.1%)<sup>‡</sup> | - | - | - | - |  |
| Baseline MoCA score<sup>†</sup> | 27 ± 2 | 25 ± 3 | 29 ± 1 | 28 ± 2 | 29 ± 1 | 28 ± 2 | <0.001 |
| APOE ε4 allele carrier | 49 (25%) | 214 (26%) | - | - | - | - | 0.9 |
| Years in study | 8 ± 4 | 5 ± 4 | 12 ± 2 | 12 ± 2 | 10 ± 4 | 10 ± 4 | <0.001 |
| Dementia at baseline | - | 43 (5%) | - | 0 | - | 0 |  |
| Dementia during follow-up | - | 108 (12%) | - | 23 (13%) | - | 37 (14%) |  |
| Dementia-free | - | 747 (84%) | - | 158 (87%) | - | 226 (86%) |  |
| No. with longitudinal sample(s)* | 44 (21%) | 373 (42%) | 78 (89%) | 181 (100%) | 131 (100%) | 259 (99%) |  |
| Time between samples (years) | 4.1 ± 2 | 3.2 ± 0.2 | 4.4 ± 1.8 | 4.5 ± 1.1 | 2.7 ± 1.1 | 3.0 ± 0.6 | <0.001 |

Continues variables are reported as mean ± standard deviation. P-293 = Project 293. P-277 = Project 277.
\* OPDC Discovery cohort and PPMI cohort P-277 participants had only one longitudinal sample. For PPMI cohort P-293, participants had either two (PD = 25, HC = 2) or three (PD = 156, HC = 76) longitudinal samples. For PPMI cohort P-277, participants had either two (PD = 259, HC = 107) or four (PD = 0, HC = 24) longitudinal samples.
<sup>†</sup> Montreal Cognitive Assessment, adjusted for years of education.
<sup>‡</sup> Participant wheelchair-bound due to paralysis unrelated to PD. Participant excluded from Chi-squared test

### Time-to-event-analyses: Incident dementia

In the OPDC Discovery cohort, 108 participants developed incident dementia during the course of follow-up. Of the 5,408 proteins investigated in the time-to-event analysis, only one, glial fibrillary acidic protein (GFAP), was significantly associated with incident dementia in PD (HR = 2.43; 95% CI: 1.79–3.30; *p-adjust* =1.35 × 10^-4^) (**Figure 1**). Incident dementia was also associated with age and sex, and there was no evidence of violation of the proportional hazards assumption (global PH test *p* = 0.85). To ensure that the observed association was not driven by serum GFAP acting as a proxy for baseline cognitive performance or genetic risk, sensitivity analyses were conducted using additional Cox models that included APOE ε4 carrier status and baseline MoCA score as covariates. In both models, the association between baseline serum GFAP and incident dementia remained significant. Respectively. The association between baseline serum GFAP and incident dementia remained (**Supplementary Table 2**). A Kaplan–Meier plot stratified by GFAP tertiles demonstrated a dose–response relationship, with progressively higher GFAP levels associated with an increased risk of incident dementia over time (log-rank test *p* = 4.67 × 10^-19^) (**Figure 1**). Sensitivity analyses using slightly alternative definitions of dementia yielded consistent results (**Supplementary Figure 7**).

**Figure 1.**
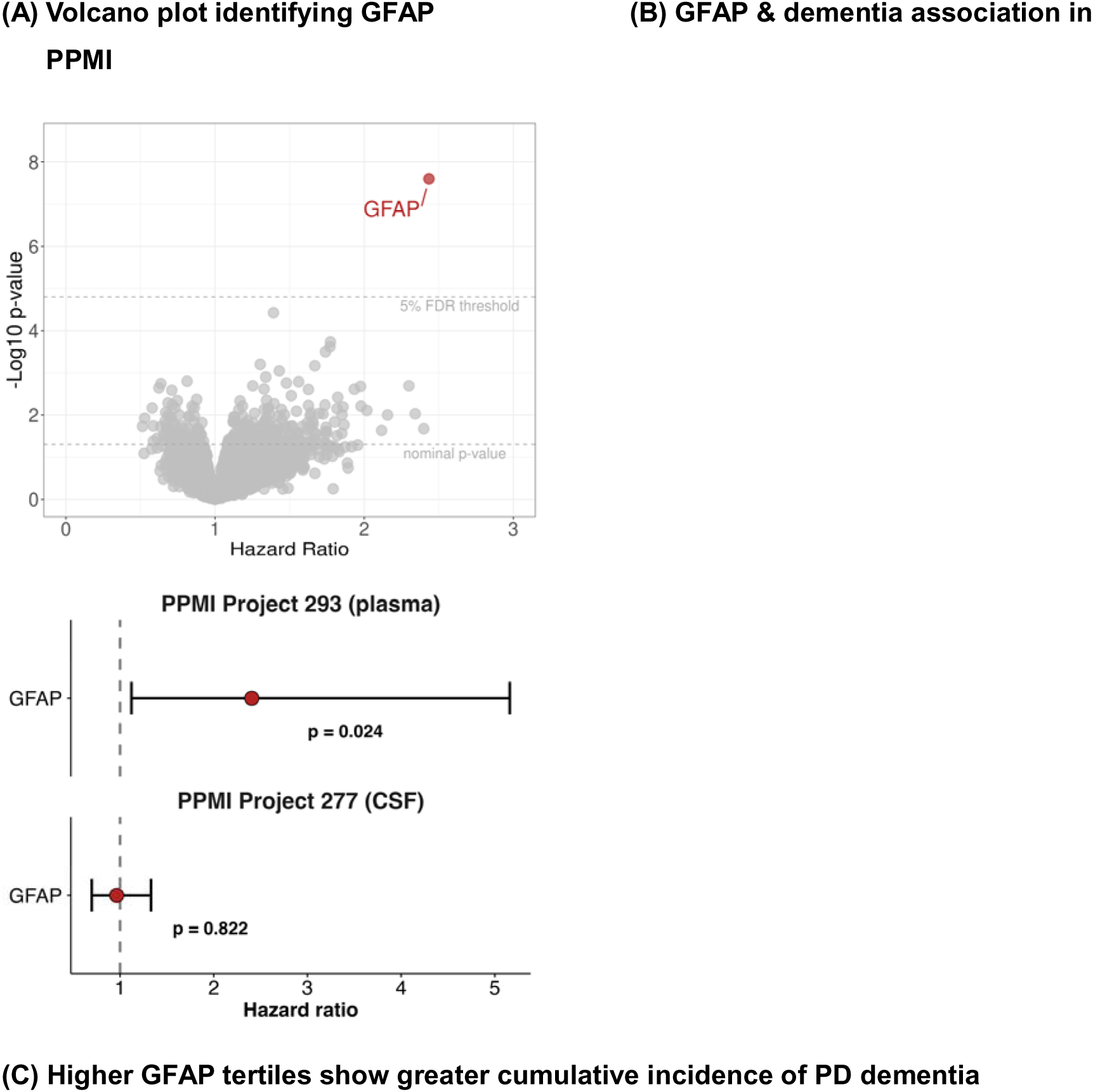
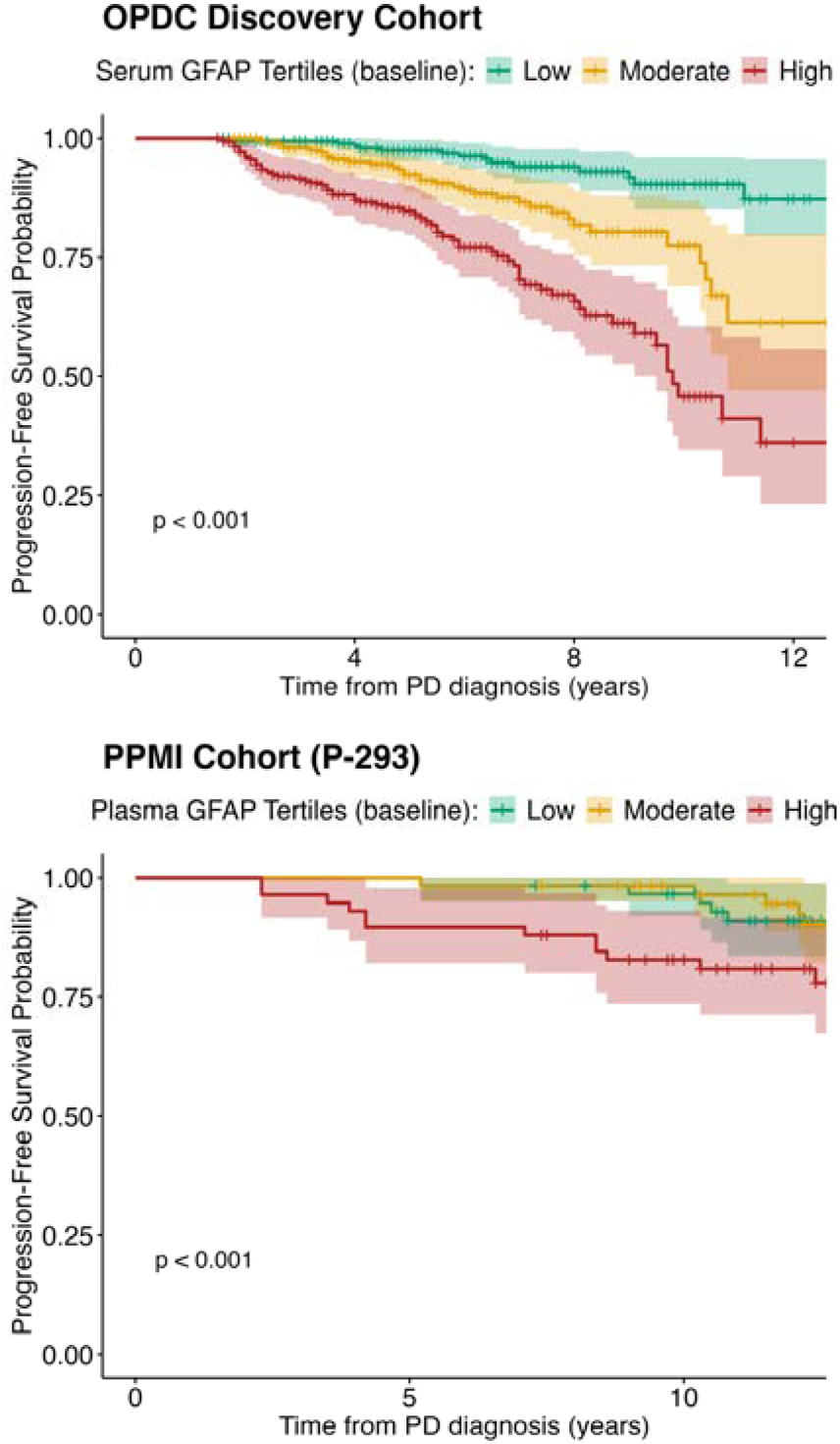
Protein Associations with incident PD dementia in the OPDC Discovery cohort. **(A)** Multivariate, delayed entry, Cox hazard regression analyses of 5408 proteins. PD participants included with at least one follow-up visit and no dementia at baseline (n = 716). Covariates: age, sex, BMI, sample age. All p-values FDR-corrected. **(B)** Forest plot showing results from time-to-event analyses in PPMI Projects 293 and 277. In both multivariable Cox regression models were used, adjusting for age, sex and BMI. **(C)** Kaplan–Meier curves showing progression to dementia stratified by baseline GFAP tertiles in the OPDC Discovery and PPMI cohorts, with shaded 95% confidence intervals and p-values from Cox proportional hazards models comparing groups. In the OPDC Discovery cohort, GFAP tertiles were defined as T1: −0.81 to −0.15 NPX, T2: −0.15 to 0.42 NPX, and T3: 0.42 to 2.78 NPX. In the PPMI cohort, tertiles were defined as T1: −1.99 to −0.52 NPX, T2: −0.52 to 0.00 NPX, and T3: 0.00 to 1.69 NPX. P-values derived from likelihood ratio tests of Cox proportional hazards models comparing dementia risk across GFAP tertiles, adjusted for age, sex and BMI and accounting for delayed entry.

In PPMI Project 293 (plasma), 23 participants developed incident dementia. The time-to-event analysis indicated that GFAP levels were significantly associated with an increased hazard of incident dementia (HR = 2.42; 95% CI: 1.12–5.22; *p* = 0.02), and the same dose-response relationship was reflected in the Kaplan-Meier plot (**Figure 1**). Sensitivity analyses using slightly different definitions of dementia yielded consistent results (**Supplementary Figure 8**). A more parsimonious model excluding BMI to reduce the risk of overfitting, given the low number of events, produced similar results (**Supplementary Table 3**). The same analysis was performed in PPMI Project 277 (CSF), where 37 participants developed incident dementia; however, no significant association between GFAP and incident dementia was observed (HR = 0.96; 95% CI: 0.66–1.39; *p* = 0.82) (**Figure 1**). Sensitivity analyses using slightly different definitions of dementia yielded consistent results (**Supplementary Figure 9**). Among 208 PPMI participants, 229 paired plasma and CSF GFAP measurements were obtained at the same visit (PD, n = 146 pairs; healthy controls, n = 83 pairs). In healthy controls, plasma and CSF GFAP levels were weakly but significantly correlated (r = 0.33, *p* = 0.002). The correlation in PD was very weak and not statistically significant (r = 0.16, *p* = 0.06) (**Supplementary Figure 10**).

Next, PD cases from the OPDC Discovery cohort were stratified into five contrasting groups: (i) PD dementia at baseline; (ii) PD dementia within 5 years of enrolment; (iii) PD dementia within 5-10 years of enrolment; (iv) PD with normal cognition after ≥5 years; and (v) PD normal cognition after 10+ years. Normal cognition was defined as an education-adjusted MoCA of ≥26/30. Compared with healthy controls, serum GFAP concentrations were higher in participants with PD incident dementia at any time point (baseline: β = 0.44, 95% CI: 0.22–0.65, *p-adjust* = 1.09 × 10^-4^; within 5 years: β = 0.39, 95% CI: 0.22–0.56, *p-adjust* = 4.30 × 10^-5^; after 5-10 years: β = 0.46, 95% CI: 0.19–0.71, *p-adjust* = 8.54 × 10^-4^). In contrast, serum GFAP levels in participants with PD who remained cognitively normal after ≥5 or ≥10 years were similar to those in healthy controls (after ≥5 years: β = 0.05, 95% CI: - 0.08–0.19, *p-adjust* = 0.47, β = 0.14, 95% CI:-0.06–0.34, *p-adjust* = 0.22) (**Figure 2**).

**Figure 2.**
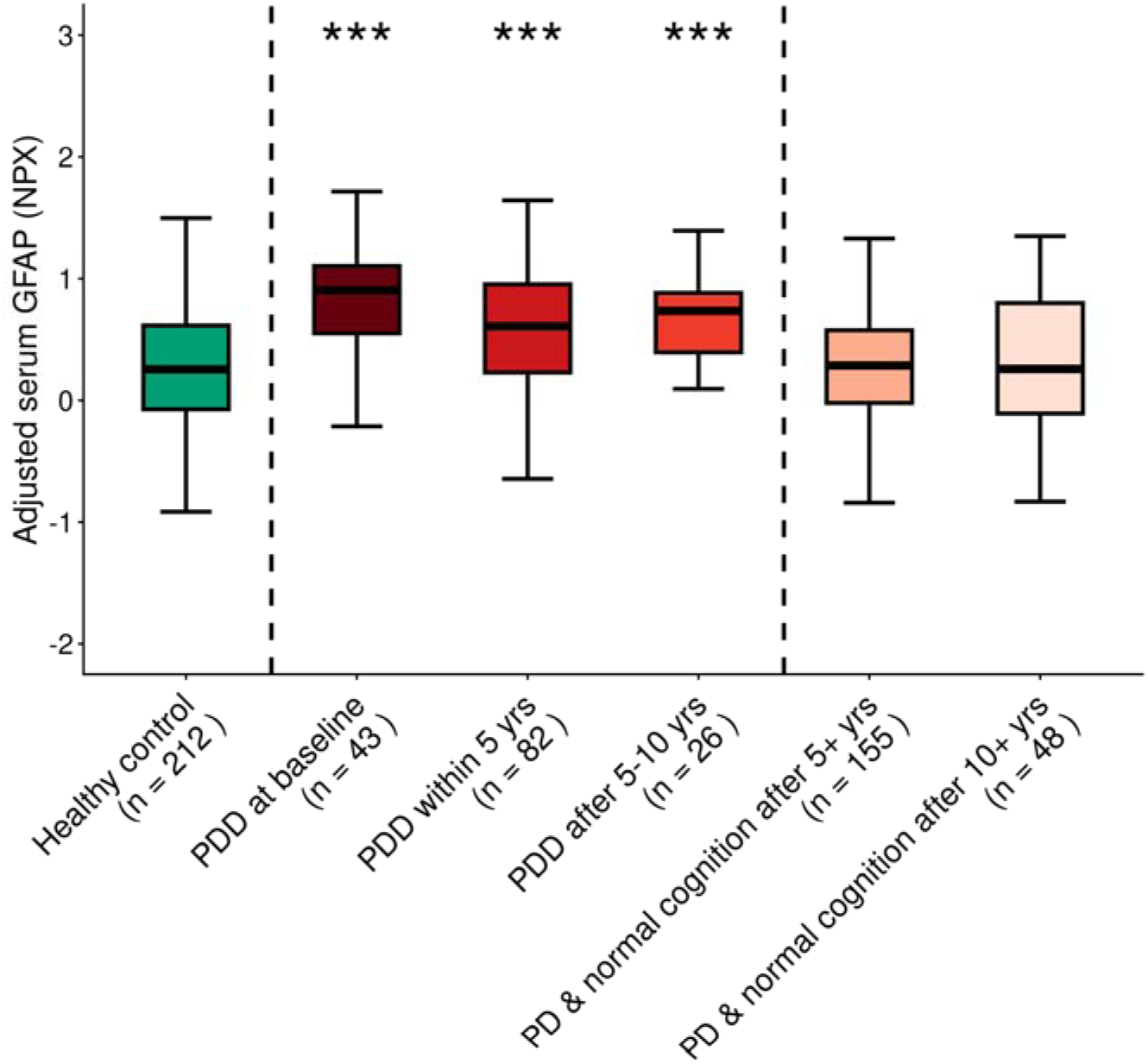
**Serum GFAP in baseline and incident dementia (OPDC Discovery Cohort)**Covariate-adjusted serum GFAP NPX displayed. PDD = Parkinson’s disease dementia. Normal cognition = MoCA score ≥26/30. Associations between selected groups and GFAP were assessed using linear regression models, adjusted for age, sex, disease duration, BMI, APOE ε4 carrier status and sample age. *** p < 0.05, ** p < 0.01, * p < 0.001. All p-values FDR-corrected.

To assess longitudinal changes in serum GFAP, repeated samples from a subset of participants in each dataset were analysed. In the OPDC Discovery cohort, serum GFAP increased over time (β = 0.11, 95% CI 0.05–0.18, *p* = 4.85 × 10^-4^). However, there was no evidence that the rate of change differed significantly between healthy controls, PD cases with or who developed dementia, and PD cases without dementia (time × PD with/incident dementia: β =-0.06, 95% CI-0.14–0.01, *p* = 0.11; time × PD without dementia: β =-0.05, 95% CI-0.12–0.02, *p* = 0.14). A further analysis focusing on more contrasting groups (PD cases with early dementia versus those with normal cognition after ≥5 years) yielded similar results, with no significant change in serum GFAP levels observed over time in healthy controls (β =-0.21, 95% CI-0.44–0.03, *p* = 0.09), PD cases with dementia at enrolment (β =-0.07, 95% CI-0.32–0.20, *p* = 0.62), PD cases who developed dementia within 5 years (β = - 0.005, 95% CI-0.22–0.22, *p* = 0.96), or PD cases without dementia after 5 years (β =-0.064, 95% CI-0.23–0.10, *p* = 0.43) (**Figure 3**). In the PPMI cohort, plasma GFAP increased over time (β = 0.056, 95% CI 0.04–0.07; *p* = 2.88 × 10^-13^). However, there was no evidence that the rate of change differed between PD cases who developed dementia and those without dementia (time × PD incident dementia: β = 0.02, 95% CI-0.01–0.05, *p* = 0.18; time × PD without dementia: β = 0.004, 95% CI-0.01–0.02, p = 0.67). Similarly, CSF GFAP increased over time (β = 0.13, 95% CI 0.09–0.18, *p* = 2.12 × 10^-8^), with no evidence of differential longitudinal change between groups (time × PD incident dementia: β =-0.01, 95% CI-0.12– 0.10; p = 0.83; time × PD without dementia: β = −0.05, 95% CI-0.11–0.01, *p* = 0.12) (**Figure 3**).

**Figure 3.**
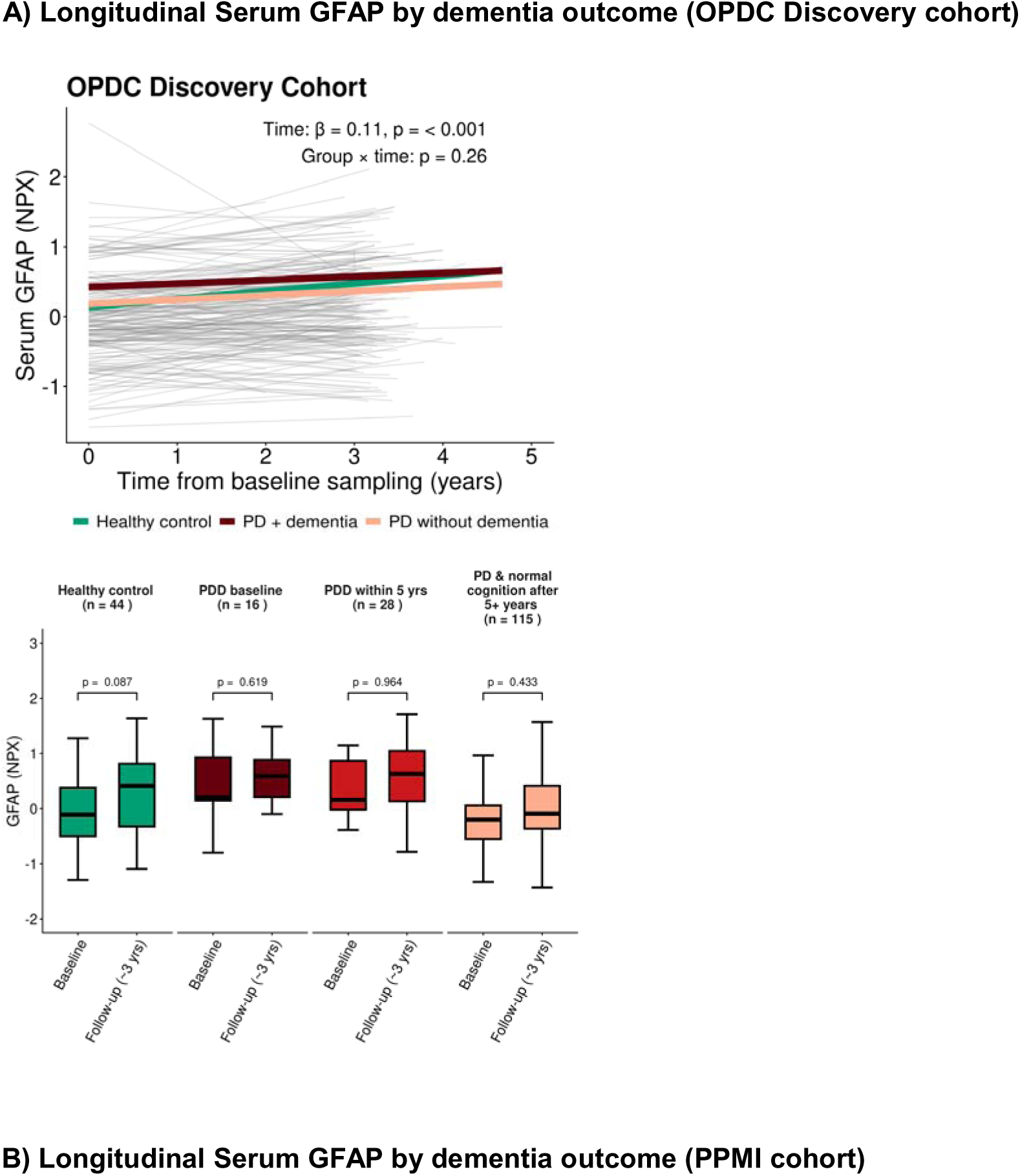
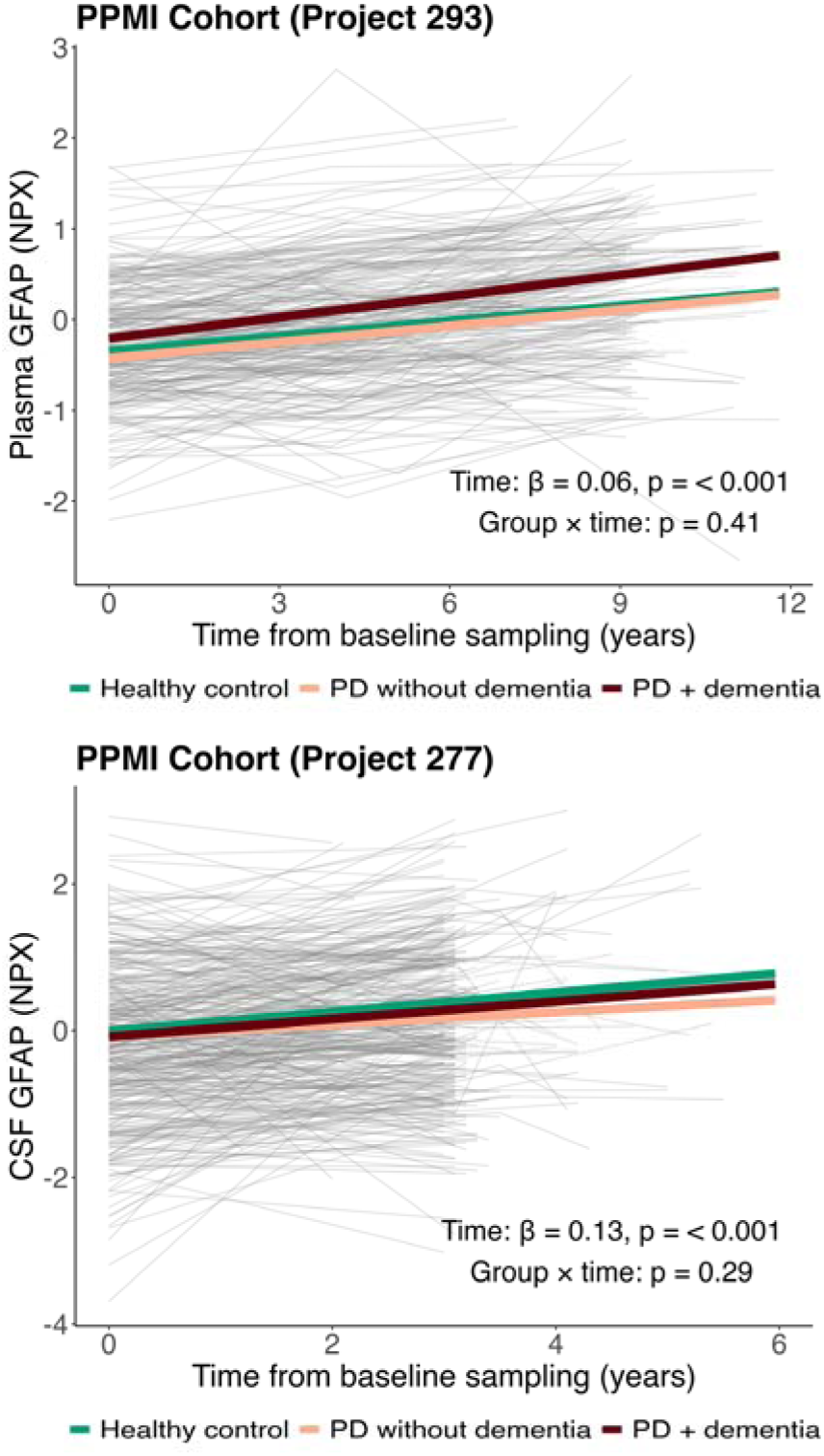
Serum GFAP in baseline and incident dementia. **(A)** Raw GFAP NPX displayed. PD without dementia: no baseline or future dementia. PD + dementia: baseline and/or incident dementia. Time is measured from the baseline visit. Associations between cognitive status and GFAP levels were assessed using linear mixed-effects models. Additional fixed effects included age, sex, disease duration, BMI, APOE ε*4 carrier status at baseline, and* sample age. A random intercept was included for participant ID. For the left-hand plot, a global ANOVA test of the group × time interaction term was used to assess whether GFAP trajectories differed between groups. **(B)** Raw GFAP NPX displayed. PD without dementia: no baseline or future dementia. PD + dementia: baseline and/or incident dementia. Individual trajectories of GFAP levels across 2-4 time points are shown, along with model-predicted trajectories. GFAP predictions are based on a linear mixed-effects model with additional fixed effects for baseline age, sex, disease duration, and BMI, and a random intercept for participant ID. A global ANOVA test of the group × time interaction term was used to assess whether GFAP trajectories differed between groups.

### GFAP and clinical variables

Serum GFAP levels were associated with several core clinical and demographic variables in both arms of the ODPC Discovery cohort. Increasing age showed a positive association with GFAP (HC: β = 0.05, 95% CI 0.04–0.06, *p* = 5.58 × 10^-14^; PD: β = 0.04, 95% CI 0.04–0.05, *p* = 3.05 × 10^-78^) while males exhibited lower levels than females (HC: β = 0.31, 95% CI 0.16–0.47, *p* = 8.89 × 10^-5^; PD: β = 0.35, 95% CI 0.27–0.43, *p* = 6.93×10^-17^). Higher BMI was associated with lower GFAP levels (HC: β =-0.02, 95% CI-0.04−0.004, *p* = 0.02; PD: β = - 0.02, 95% CI-0.026−-0.009, *p* = 4.09 × 10^-5^). APOE ε4 carrier status was not significantly associated with GFAP in either group (HC ε4−/− vs ε4+/−: β =-0.08, 95% CI-0.26−0.10; *p-adjust* = 0.70; PD ε4−/− vs ε4+/−: β =-0.090, 95% CI-0.18−-0.002; *p-adjust* = 0.18). Serum GFAP was significantly higher in the PD group of the Discovery cohort (β = 0.13, 95% CI 0.04−0.22, *p* = 0.003), but neither plasma nor CSF GFAP was significantly associated with PD in the PPMI projects (PPMI Project 293: β =-0.01, 95% CI-0.15−0.13, *p* = 0.87; PPMI Project 277: β =-0.02, 95% CI-0.22−0.18, *p* = 0.82) (**Supplementary Figure 9**). With respect to cognitive performance and motor severity, higher baseline serum GFAP levels were associated with lower MoCA scores at baseline (β = −0.48, 95% CI-0.87−-0.08, *p* = 0.02) and mildly worse MDS-UPDRS III scores at baseline consistent with chance (β = 1.27, 95% CI-0.04−2.58, *p* = 0.06). In longitudinal analyses, higher baseline GFAP was associated with a greater decline in MoCA over follow-up (GFAP × time: β =-0.34, 95% CI - 0.50−-0.18, *p* = 2.93 × 10^-5^). There was no statistically significant association between baseline serum GFAP and change in MDS-UPDRS III scores over time though the coefficient was positive (GFAP × time: β = 0.65, 95% CI-0.03−1.32, *p* = 0.06) (**Figure 4**).

**Figure 4.**
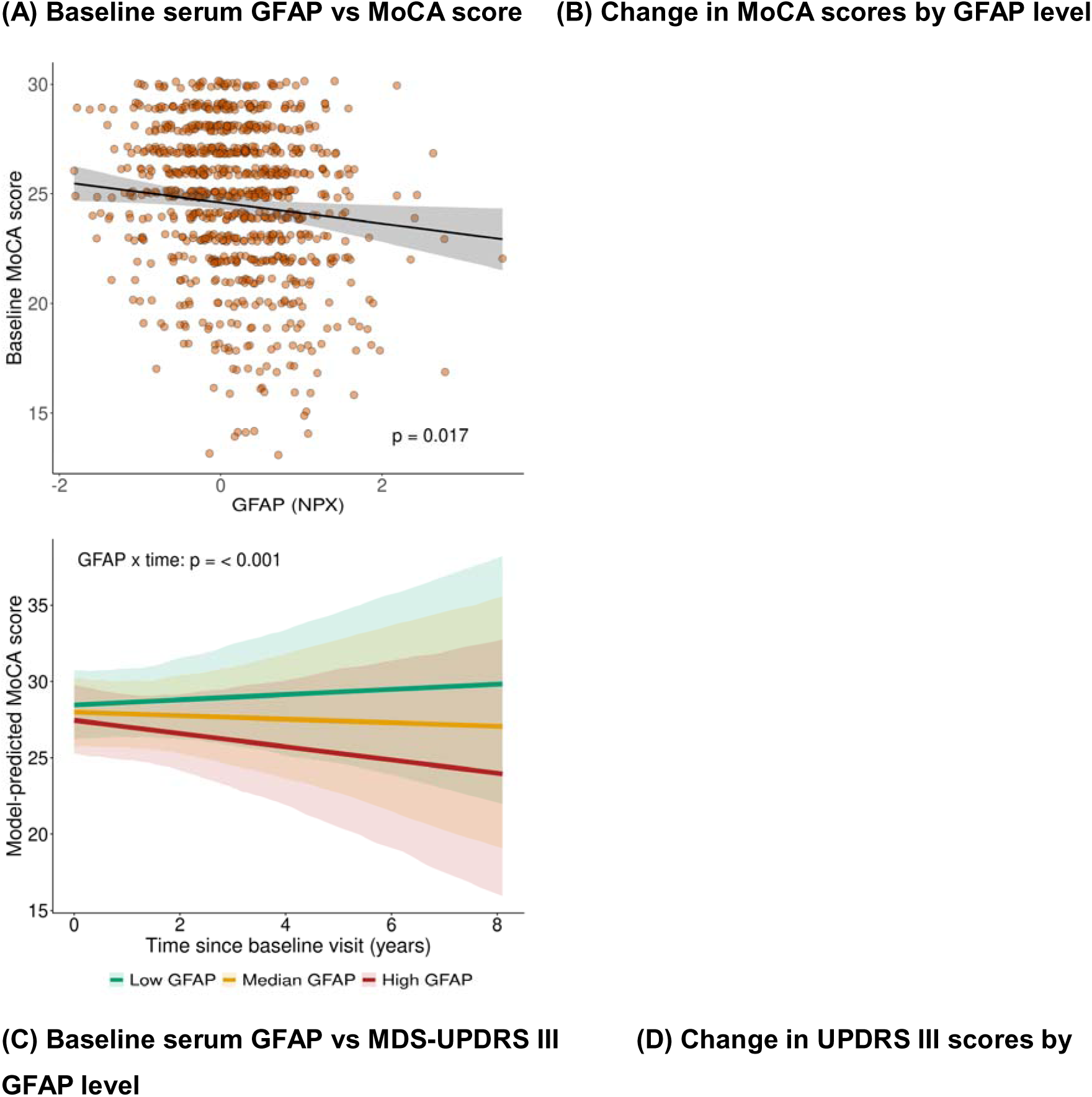
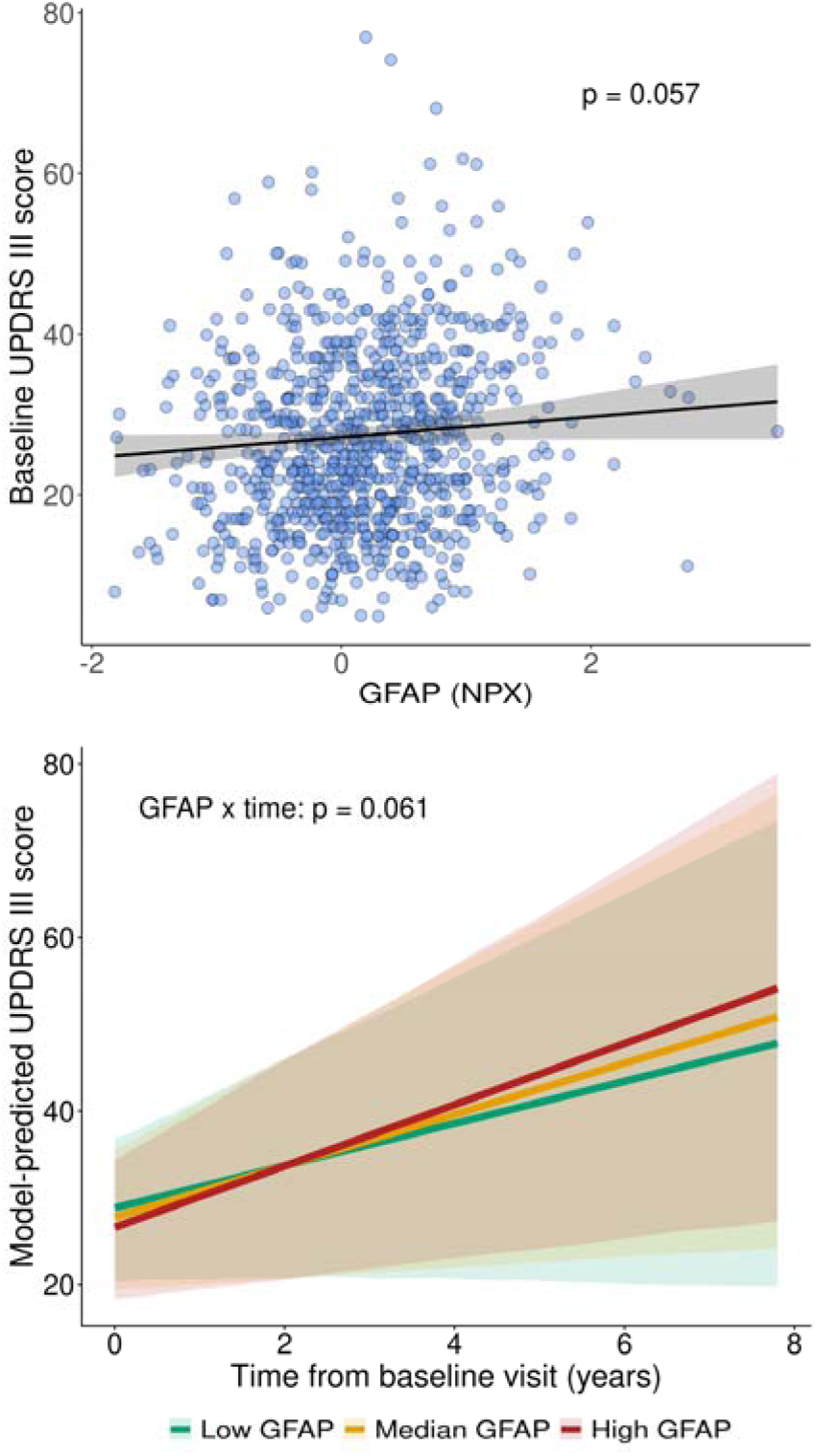
Associations between baseline serum GFAP and clinical variables. **(A)** Raw GFAP NPX displayed. Linear regression employed to assess association of GFAP with education-adjusted MoCA score at baseline (PD cases only, n = 883). Covariates: age, sex, BMI, APOE ε4 carrier status and sample age. **(B)** Population-level predicted MoCA trajectories from a linear mixed-effects model including a baseline GFAP × time interaction (PD cases only, n = 830). Fixed effects: disease duration, age, sex and sample age at baseline. Random intercept: participant ID. Low GFAP = 10^th^ percentile, Median GFAP = 50^th^ percentile, High GFAP = 90^th^ percentile. **(C)** Raw GFAP NPX displayed. Linear regression model assessing association of GFAP with MDS-UPDRS III at baseline (PD cases only, n = 875). Covariates: age, sex, BMI, APOE ε4 carrier status and sample age. **(D)** Population-level predicted MoCA trajectories from a linear mixed-effects model including a baseline GFAP × time interaction (PD cases only, n = 827). Fixed effects: disease duration, age, sex and sample age at baseline. Random intercept: participant ID. Low GFAP = 10^th^ percentile (NPX = - 0.77), Median GFAP = 50^th^ percentile (NPX = 0.05), High GFAP = 90^th^ percentile (NPX = 0.96).

## Discussion

In this large, longitudinal proteomic study across two independent cohorts, we identify circulating GFAP as a robust and reproducible predictor of future dementia in PD. Among more than 5,400 proteins interrogated in the OPDC Discovery cohort, GFAP emerged as the sole biomarker significantly associated with incident dementia, with consistent effect sizes observed in both the OPDC Discovery cohort and the independent PPMI plasma dataset. These findings suggest that peripheral GFAP captures a key biological process present many years before the onset of cognitive decline in PD and represents a potentially powerful prognostic biomarker early in the disease course.

GFAP is an intermediate filament protein expressed primarily in astrocytes and is widely regarded as a marker of astroglial activation and injury.^31–33^ In neurodegenerative disease broadly, elevated plasma GFAP has been identified as a predictor of future all-cause dementia in several large prospective cohort studies.^34–36^ In Alzheimer’s disease (AD) GFAP is an established biomarker, with levels elevated early in the disease course and higher concentrations associated with an increased risk of cognitive decline.^37–40^ Notably, plasma GFAP outperforms CSF GFAP in distinguishing AD cases from healthy controls and shows stronger associations with core CSF biomarkers of AD pathology.^41^ In dementia with Lewy bodies (DLB) and Parkinson’s disease dementia (PDD), emerging evidence indicates that both CSF and plasma GFAP levels are elevated, with higher plasma baseline concentrations associated with more rapid disease progression.^42–44^ In PD, blood-based GFAP levels show only modest or inconsistent separation between cases and controls across studies.^45–48^ When phenotypically stratified however, higher levels have been observed in patients with postural instability and gait disorder (PIGD) compared to tremor-dominant (TD) PD and those with a higher non-motor symptom burden.^49–51^ Adding to this, a recent report demonstrated higher plasma GFAP levels in PD patients with dementia compared to those without, while another study showed that elevated CSF GFAP was associated with greater cognitive decline.^22,23^ In light of the fact that there is a known association between the PIGD motor-subtype and greater non-motor symptom burden and cognitive decline in PD^20,21,52^ and taking into account the broader evidence linking elevated plasma GFAP to dementia, current evidence would suggest that elevated plasma GFAP is a marker of cognitive decline in PD. Our findings build on this existing literature and demonstrates that circulating GFAP has clear prognostic value in PD. Higher baseline GFAP levels were significantly associated with both an increased hazard of incident dementia and a steeper trajectory of cognitive decline, as measured by MoCA at baseline and over time. Of particular note, PD patients who maintained preserved cognition (defined as a MoCA score of ≥ 26/30) after ≥5 or ≥10 years showed no discernible difference in serum GFAP levels compared with healthy controls at baseline. By contrast, MDS-UPDRS III scores showed only a weak association with GFAP levels at baseline and longitudinally supporting serum GFAP as a marker of cognitive rather than motor decline.

The observed dose-response relationship across GFAP tertiles supports a biologically meaningful gradient of dementia risk, suggesting that GFAP may signal the underlying disease process driving cognitive decline in PD. Growing evidence suggests that concomitant AD pathology in both manifest and prodromal PD predicts future dementia.^17–19,53^ Post-mortem studies of PD and AD have shown higher plasma GFAP levels in individuals with a greater burden of AD pathology, while studies across the AD continuum demonstrate that GFAP is positively associated with the extent of amyloid-β pathology.^41,54–57^ Our findings strengthen the evidence that GFAP predicts future dementia in PD and indirectly support the hypothesis that elevated GFAP reflects underlying concomitant AD pathology many years before cognitive decline manifests.

Longitudinal analyses across both the OPDC Discovery and PPMI cohorts showed a clear increase in both blood-based and CSF GFAP levels over time. A notable difference was observed between the cross-sectional age coefficient (β ≈ 0.05) and the longitudinal time coefficient (β ≈ 0.11). The cross-sectional association with age may be attenuated by measurement variability, whereas longitudinal models, which capture within-individual change, are less susceptible to such bias. Additional unmeasured factors may contribute to this discrepancy; however, PD or dementia risk are unlikely to explain it, as there was no evidence that GFAP trajectories differed between healthy controls, PD patients with or who developed dementia, and PD patients without dementia. This suggests that GFAP does not act as a dynamic marker of cognitive decline in PD. Instead, these findings support the interpretation that GFAP reflects an early, underlying biological vulnerability or subclinical process, consistent with the broader PD aetiological framework in which preclinical pathological changes precede symptom onset by many years or decades.^58^

Finally, this study demonstrates a notable discrepancy between plasma and CSF GFAP, with only peripheral – but not CSF – GFAP showing a significant association with incident dementia, a weak correlation between plasma and CSF GFAP in healthy controls and no significant correlation in PD in the PPMI cohort. While at its surface counterintuitive, this aligns with what has been observed in AD research, where prior work has demonstrated stronger diagnostic and prognostic performance for plasma compared to CSF GFAP.^41,56^ Although it remains poorly understood, several mechanisms have been proposed to explain this divergence. Plasma GFAP is thought to better reflect global astroglial activation and early amyloid-related pathology, potentially amplified by blood–brain barrier dysfunction and glymphatic clearance pathways that facilitate the release of GFAP into the circulation. In contrast, CSF GFAP may capture more localized or compartmentalised astrocytic responses within the central nervous system, which may be less sensitive to early or diffuse pathological processes.^41,56,59^

## Limitations

Several limitations should be considered when interpreting these findings. First, the definition of dementia, while based on a structured and clinically informed algorithm incorporating MoCA thresholds and MDS-UPDRS items alongside clinical diagnosis, may still be susceptible to misclassification, particularly in early or borderline cases where cognitive impairment is subtle or fluctuating. Second, the relatively small number of incident dementia events in the PPMI cohort projects increases the risk of overfitting and imprecise effect estimates when implementing Cox models. This is particularly relevant for the null findings observed in the CSF analysis, which may reflect limited power rather than a true absence of association. Third, a limitation stems from the use of NPX units to quantify protein levels, which provide relative rather than absolute measurements. Consequently, observed differences in protein expression are intrinsically tied to the specific experimental conditions of this study, limiting generalisability and making direct comparisons with other studies challenging in the absence of bridging samples. Finally, although key confounders were adjusted for, residual confounding cannot be excluded, and mechanistic inferences remain limited in the absence of complementary imaging or neuropathological data.

## Conclusion

In this large, prospective longitudinal study across two independent cohorts, we identify circulating GFAP as a robust predictor of future dementia in Parkinson’s disease. Elevated baseline serum or plasma GFAP – but not CSF – was associated with an increased risk of incident dementia and accelerated cognitive decline, while demonstrating no significant relationship with motor scores, supporting its specificity as a marker of future cognitive impairment. Longitudinal analyses indicate that while GFAP levels increase over time in both blood and CSF, these trajectories do not differ between healthy controls, PD patients with or who developed dementia, and those without dementia. This suggests that GFAP is not a dynamic marker of cognitive progression, but instead reflects a slowly evolving biological process, indexing early, subclinical vulnerability. Together, these findings position peripheral GFAP as a promising, accessible biomarker for early risk stratification in PD. Future studies integrating GFAP with complementary biomarkers and imaging modalities will be essential to refine its clinical utility and to further elucidate the underlying mechanisms linking astroglial activation to cognitive decline in PD.

## Data Availability

Data from the OPDC Discovery cohort are not publicly available because of participant confidentiality, consent restrictions, and the proprietary nature of the data. Deidentified data may be made available from the corresponding author upon reasonable request, subject to appropriate approvals.

## Acknowledgements

The authors wish to express their sincere gratitude to the participants of the Oxford Parkinson’s Disease Centre (OPDC) Discovery Cohort and their families for their invaluable contributions. We also extend our appreciation to the personnel of the OPDC for their dedicated support. This study was funded by GSK through the Oxford-GSK Institute of Molecular and Computational Medicine (IMCM) and the Monument Trust Discovery Award from Parkinson’s UK (grant number: J-2101). PPMI – a public–private partnership – is funded by the Michael J. Fox Foundation for Parkinson’s Research and funding partners, including 4D Pharma, AbbVie, AcureX, Allergan, Amathus Therapeutics, Aligning Science Across Parkinson’s (ASAP), AskBio, Avid Radiopharmaceuticals, BIAL, BioArctic, Biogen, Biohaven, BioLegend, BlueRock Therapeutics, Bristol Myers Squibb, Calico Labs, Capsida Biotherapeutics, Celgene, Cerevel Therapeutics, Coave Therapeutics, DaCapo Brainscience, Denali, Edmond J. Safra Foundation, Eli Lilly, Gain Therapeutics, GE Healthcare, Genentech, GSK, Golub Capital, Handl Therapeutics, Insitro, Jazz Pharmaceuticals, Johnson & Johnson Innovative Medicine, Lundbeck, Merck, Meso Scale Discovery, Mission Therapeutics, Neurocrine Biosciences, Neuron23, Neuropore, Pfizer, Piramal, Prevail Therapeutics, Roche, Sanofi, Servier, Sun Pharma Advanced Research Company, Takeda, Teva, UCB, Vanqua Bio, Verily, Voyager Therapeutics, and The Weston Family Foundation. The funders played no role in study design, data collection, analysis and interpretation of data, or the writing of this manuscript.

## Artificial intelligence use

University of Oxford ChatGPT Edu workspace (version 5.4) was used to assist with debugging R code in the analytical pipeline and with rephrasing text to improve language and flow. The authors reviewed and take full responsibility for the final manuscript content.

## Data Sharing Statement

Data from the OPDC Discovery cohort are not publicly available because of participant confidentiality, consent restrictions, and the proprietary nature of the data. Deidentified data may be made available from the corresponding author upon reasonable request, subject to appropriate approvals. PPMI clinical and proteomic data are available through the data portal at: https://ida.loni.usc.edu/pages/access/studyData.jsp?categoryId=7&subCategoryId=305.

