## Supplement 1 for "Peripheral GFAP Predicts Incident Dementia in Parkinson’s Disease"

**Supplementary Materials**

**OPDC Discovery Cohort – Quality Control Summary**

**Supplementary Figure 1. Quality control flow diagram**

Total number of assays tested: 5420

2 assays (GBP1, MAP2K1) were tested in triplicate. The mean was calculated and carried forward to the downstream analysis

New total: 5416

8 assays had an Olink WARN label (APOE, BLMH, CTSD, CTSV, ELANE, GAPDH, SBSM, SERPINB4). These were excluded from downstream analysis

New total: 5408

Total number of serum samples tested: 1561

5 serum samples removed (no NPX generated – failed experiment)

New total: 1556

15 samples identified as extreme outliers and removed

New total: 1541

11 samples removed as they were not taken at an appropriate time point Two samples removed due to insufficient metadata. Six healthy controls removed that had a MoCA score of < 21 at baseline New total: 1522

**Supplementary Figure 2. Extreme outlier detection**

**Outlier Detection**

Outliers in the CSF proteomics dataset were identified using a combination of three complementary methods:

1. *Quality Control (QC) Plot Method:*

Outliers were defined as samples exceeding 5 standard deviations from the mean interquartile range (IQR) or sample median across all proteins. This approach identified 11 extreme outliers (**A**).

2. *Principal Component Analysis (PCA) – Mahalanobis Distance:*

PCA was performed on normalized protein expression (NPX) data, and Mahalanobis distance was used to identify statistical outliers with a confidence level of 99.999%, detecting 9 samples as outliers (**B**).

3. *PCA – Euclidean Distance from Centroid of Top Principal Components:*

The top 10 principal components were used to calculate each sample’s Euclidean distance from the centroid of the PD dataset. The same 9 outliers were identified as with the Mahalanobis distance approach.

**Summary:** Combining the three above techniques identified a total of 15 outliers.

**A)**

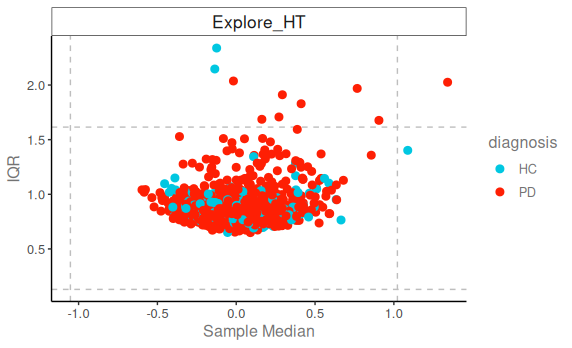

**B)**

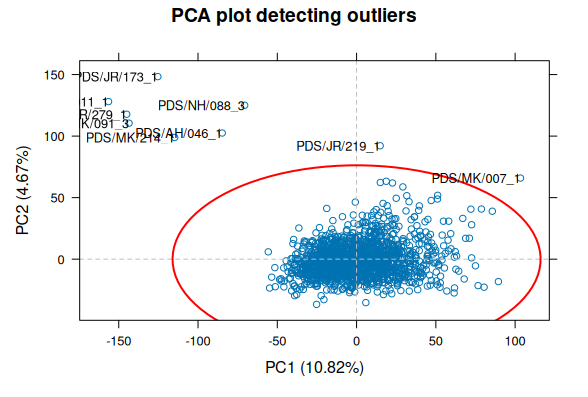

**PPMI Cohort Project 293 – Quality Control Summary**

**Supplementary Figure 3. Quality control flow diagram for PPMI Cohort projects**

PPMI Project 293	PPMI Project 277

1200 samples tested, from which the HC & PD samples with complete clinical data were selected (n = 833)

GFAP selected from >5000 protein read-outs. Olink SampleQC = FAIL: 0 cases Olink AssayQC = FAIL: 0 cases New total: 833 (393 baseline, 440 longitudinal)

1 baseline and 2 longitudinal samples identified as extreme outliers and removed. Sample paired with the 1 baseline extreme outlier also removed New total: 830 (392 baseline, 438 longitudinal)

874 samples tested, from which the HC & PD samples with complete clinical data were selected (n = 764)

GFAP selected from >5000 protein read-outs. Olink SampleQC = FAIL: 1 case Olink AssayQC = FAIL: 0 cases New total: 763 (269 baseline, 494 longitudinal)

3 longitudinal samples identified as extreme outliers and removed. New total: 760 (269 baseline, 491 longitudinal)

**Supplementary Figure 4. Outlier Plots for PPMI Cohort Projects**

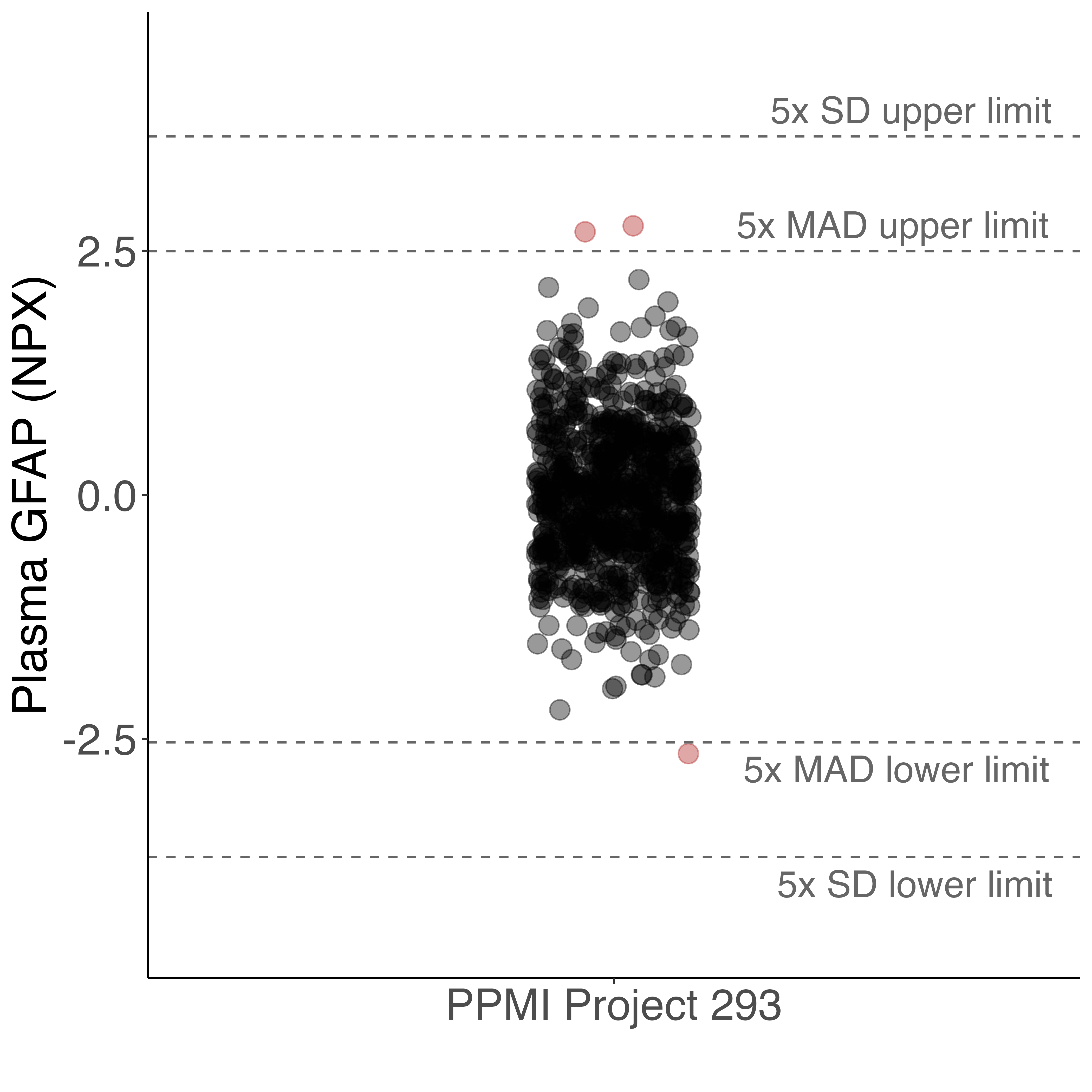

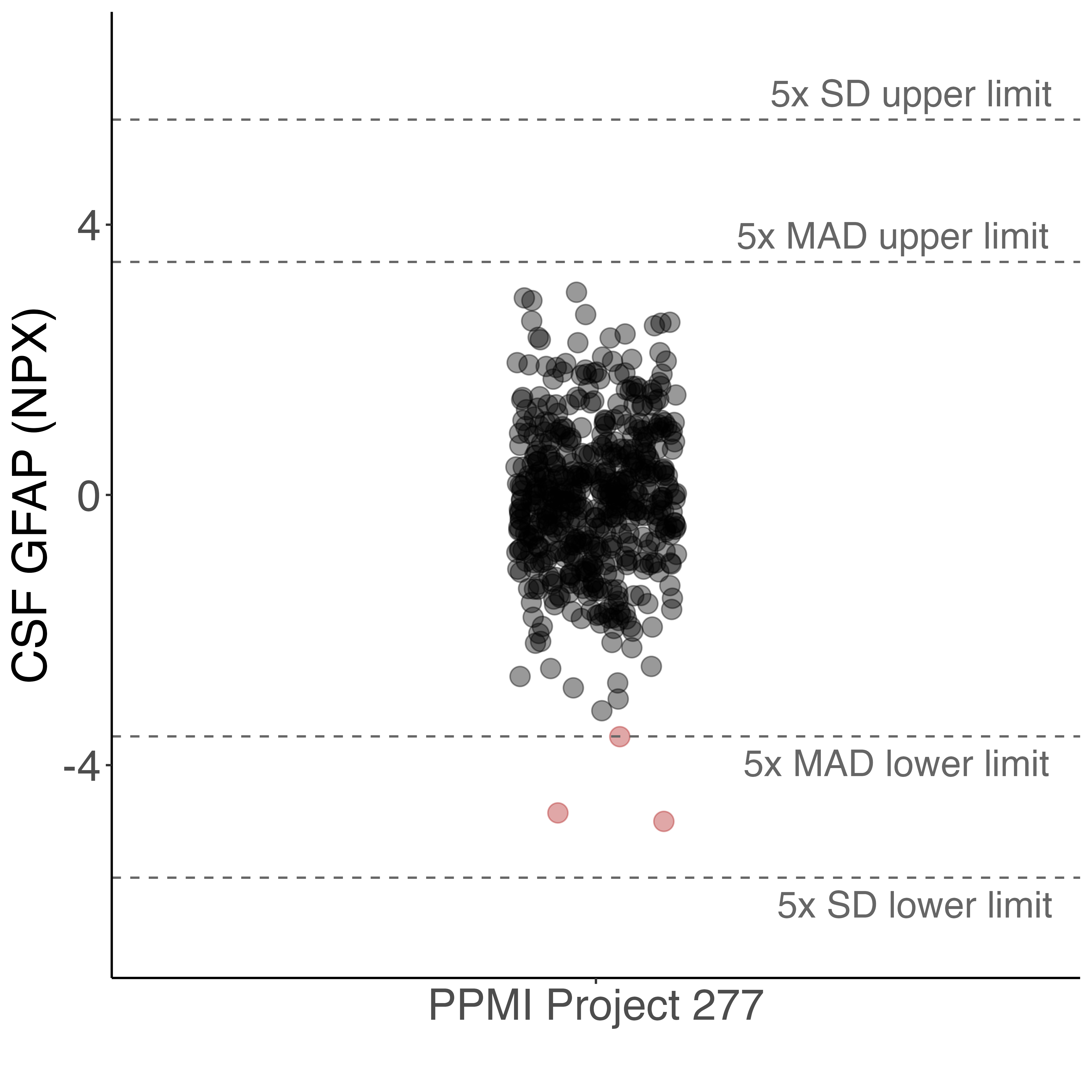

*SD = Standard deviation. MAD = Median absolute deviation. Highlighted in red are the extreme outliers. All PD and healthy control samples included in this analysis.*

**Supplementary Figure 5. Distribution of assay-level percentage below LOD**

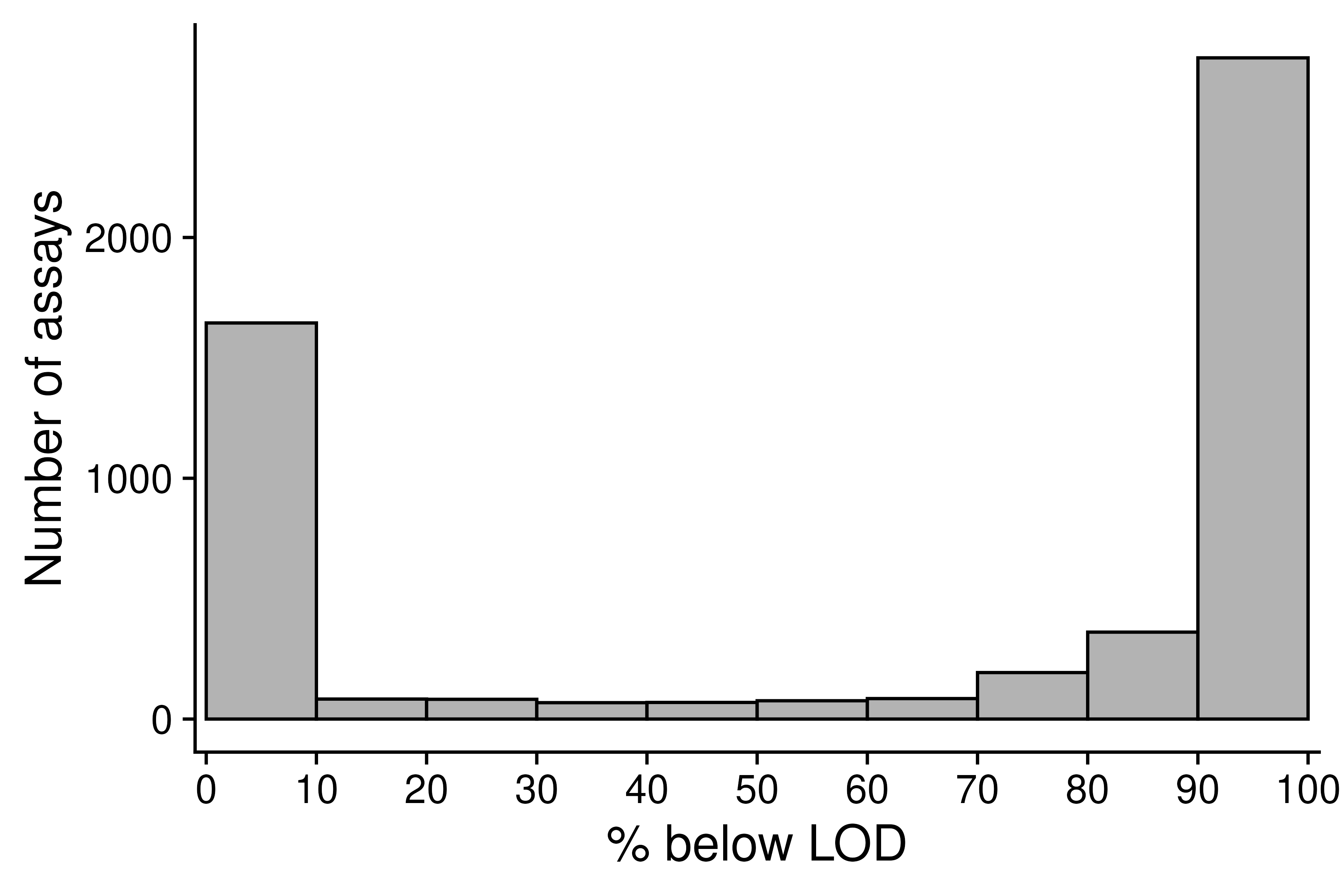

*LOD = limit of detection*

**Supplementary Table 1. Interplate coefficient of variance (CV) for GFAP in both cohorts**

| **ODPC Discovery Cohort (serum)** | |
| --- | --- |
| **Protein** | **CV** |
| Intraplate CV | 6.63 % |
| Interplate CV | 11.18 % |
| **PPMI Project 293 (plasma)** | |
| **Protein** | **CV** |
| Intraplate CV | 25.13% |
| Interplate CV | 10.86% |
| **PPMI Project 277 (CSF)** | |
| **Protein** | **CV** |
| Intraplate CV | 7.84% |
| Interplate CV | 22.47% |

**Supplementary Figure 6. Proportion of dementia classification criteria across datasets**

(A) OPDC Discovery Cohort

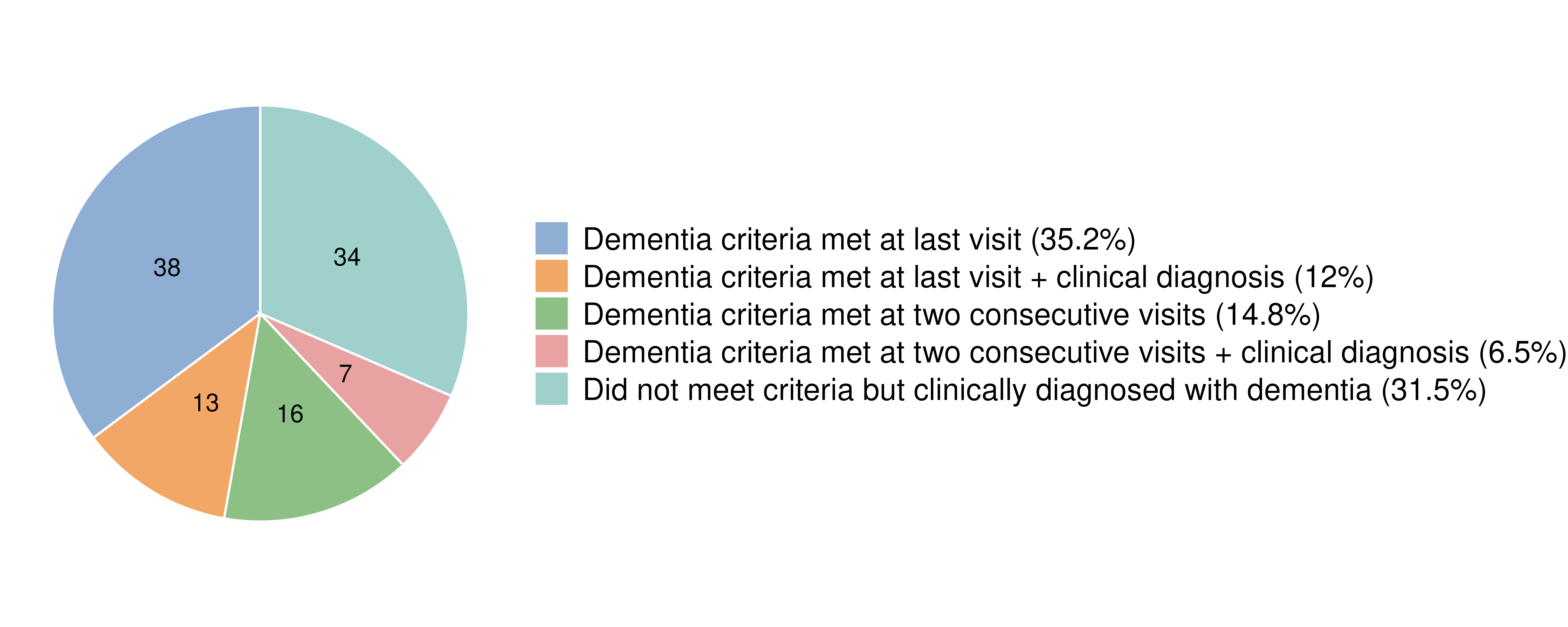

(B) PPMI Cohort – Project 293

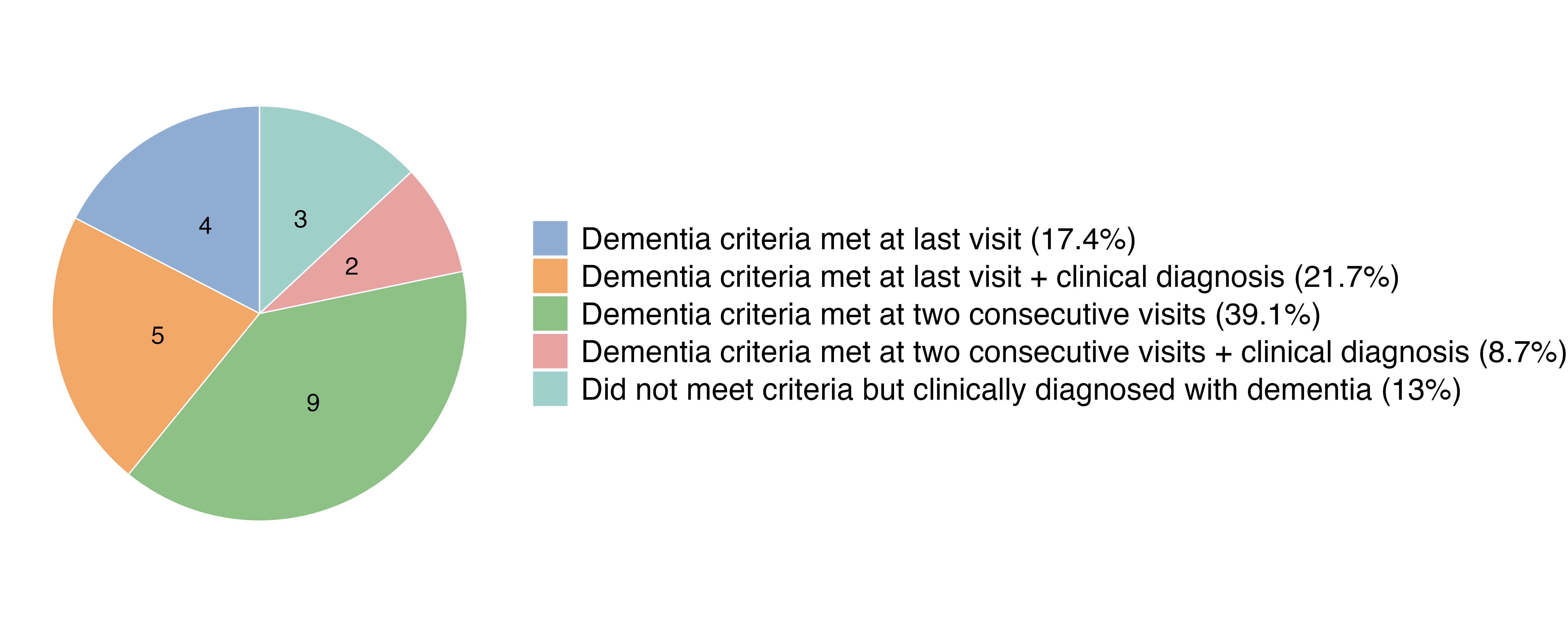

(C) PPMI Cohort – Project 277

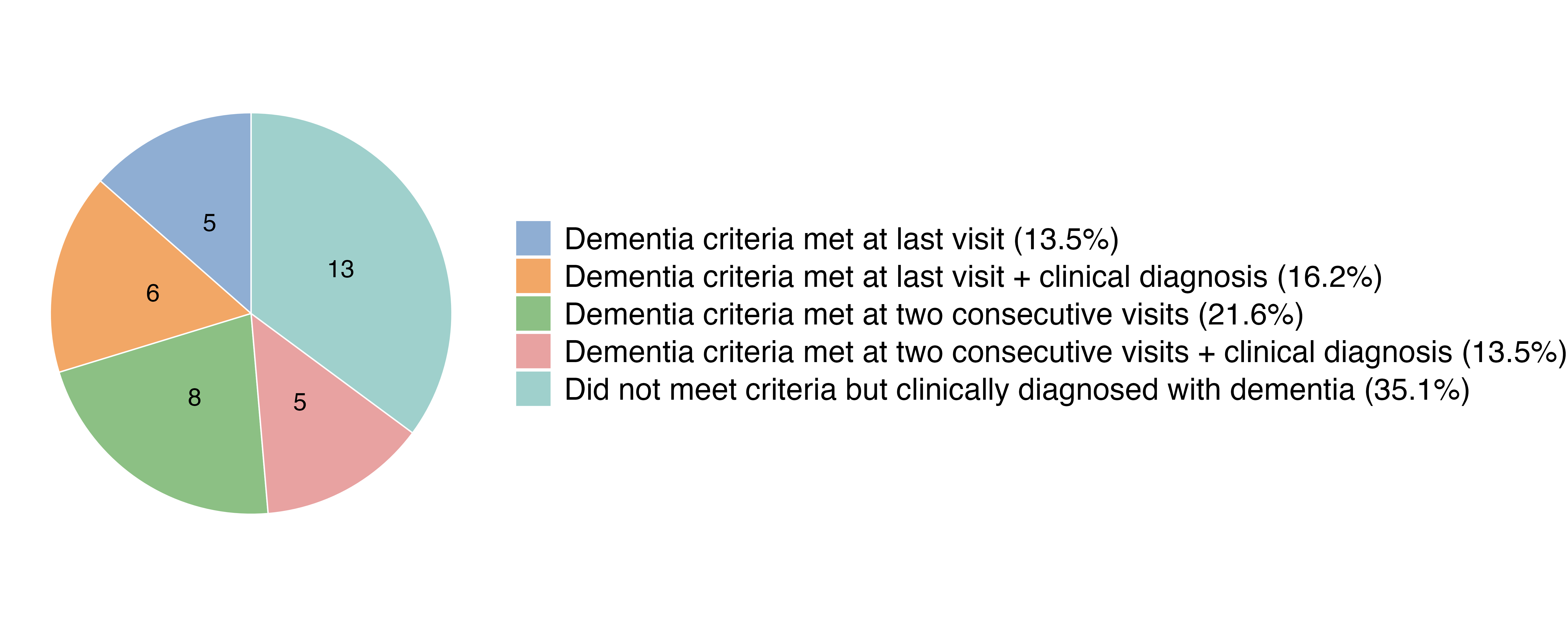

**Supplementary Table 2. Cox regression analyses in OPDC cohort (time-to-dementia)**

(A) **Baseline GFAP and Dementia Risk in PD**

Formula: *h_i_(t) = h0(t)exp(β_1_GFAP_i_ + β_2_Age_i_ + β_3_Sex_i_ + β_4_BMI_i_ + β_5_Sample_age_i_)*

| **Variable** | **β** | **Hazard Ratio (HR)** | **HR 95% CI** | **p-value** |
| --- | --- | --- | --- | --- |
| GFAP | 0.89 | 2.43 | 1.78 – 3.33 | 2.51 × 10^-8^ |
| Age | 0.07 | 1.07 | 1.03 – 1.10 | 4.73 × 10^-6^ |
| Sex | 0.62 | 1.85 | 1.20 – 2.87 | 0.005 |
| BMI | 0.03 | 1.03 | 0.98 – 1.07 | 0.24 |
| Sample age | -0.04 | 0.96 | 0.83 – 1.11 | 0.57 |

*Global PH = 0.85*

(B) **Baseline GFAP and Dementia Risk in PD + APOE ε4 status**

Formula: *h_i_(t) = h0(t)exp(β_1_GFAP_i_ + β_2_Age_i_ + β_3_Sex_i_ + β_4_BMI_i_ + β_5_Sample_age_i_ + β_6_APOE*_ε4_status*)*

| **Variable** | **β** | **Hazard Ratio (HR)** | **HR 95% CI** | **p-value** |
| --- | --- | --- | --- | --- |
| GFAP | 0.79 | 2.21 | 1.58 – 3.08 | 2.62 × 10^-6^ |
| Age | 0.07 | 1.07 | 1.04 – 1.15 | 4.78 × 10^-6^ |
| Sex | 0.41 | 1.51 | 0.96 – 2.37 | 0.074 |
| BMI | 0.03 | 1.03 | 0.98 – 1.08 | 0.203 |
| Sample age | 0.03 | 1.02 | 0.87 – 1.21 | 0.729 |
| APOE ε4 status | 0.97 | 2.63 | 1.74 – 3.97 | 3.83 × 10^-6^ |

*Global PH = 0.62*

(C) **Baseline GFAP and Dementia Risk in PD + baseline MoCA score**

Formula: *h_i_(t) = h0(t)exp(β_1_GFAP_i_ + β_2_Age_i_ + β_3_Sex_i_ + β_4_BMI_i_ + β_5_Sample_age_i_ + β_6_MoCA_i_)*

| **Variable** | **Β** | **Hazard Ratio (HR)** | **HR 95% CI** | **p-value** |
| --- | --- | --- | --- | --- |
| GFAP | 0.71 | 2.03 | 1.48 – 2.77 | 9.65 × 10^-6^ |
| Age | 0.05 | 1.05 | 1.02 – 1.08 | 0.002 |
| Sex | 0.53 | 1.70 | 1.09 – 2.65 | 0.02 |
| BMI | 0.009 | 1.009 | 0.96 – 1.06 | 0.71 |
| Sample age | -0.07 | 0.93 | 0.80 – 1.08 | 0.33 |
| MoCA score | -0.21 | 0.81 | 0.76 – 0.87 | 2.37 × 10^-10^ |

*Global PH = 0.94*

**Supplementary Table 3. Cox regression model sensitivity analysis (PPMI Project 293)**

**Given the low number of events (n = 23) and the risk of overfitting with multiple covariates, a sensitivity analysis was performed excluding BMI from the Cox model. The association between GFAP level and incident dementia remained significant.**

**Model 1**: $h(t)=h_{0}(t)\exp\left( \beta_{1}\cdot\text{GFAP}+\beta_{2}\cdot\text{Age}+\beta_{3}\cdot\text{Sex}+\beta_{4}\cdot\text{BMI} \right)$

**Model 2**: $h(t)=h_{0}(t)\exp\left( \beta_{1}\cdot\text{GFAP}+\beta_{2}\cdot\text{Age}+\beta_{3}\cdot\text{Sex} \right)$

| **Variable** | **Model 1 HR (95% CI)** | **p-value** | **Model 2 HR (95% CI)** | **p-value** |
| --- | --- | --- | --- | --- |
| GFAP | 2.41 (1.12–5.16) | 0.024 | 2.30 (1.10–4.80) | 0.026 |
| Age | 1.07 (1.01–1.13) | 0.015 | 1.07 (1.01–1.13) | 0.014 |
| Sex (male) | 3.15 (1.02–9.68) | 0.045 | 3.22 (1.05–9.87) | 0.041 |
| BMI | 1.03 (0.91–1.16) | 0.64 | — | — |

**Supplementary Figure 7. OPDC Discovery Cohort**

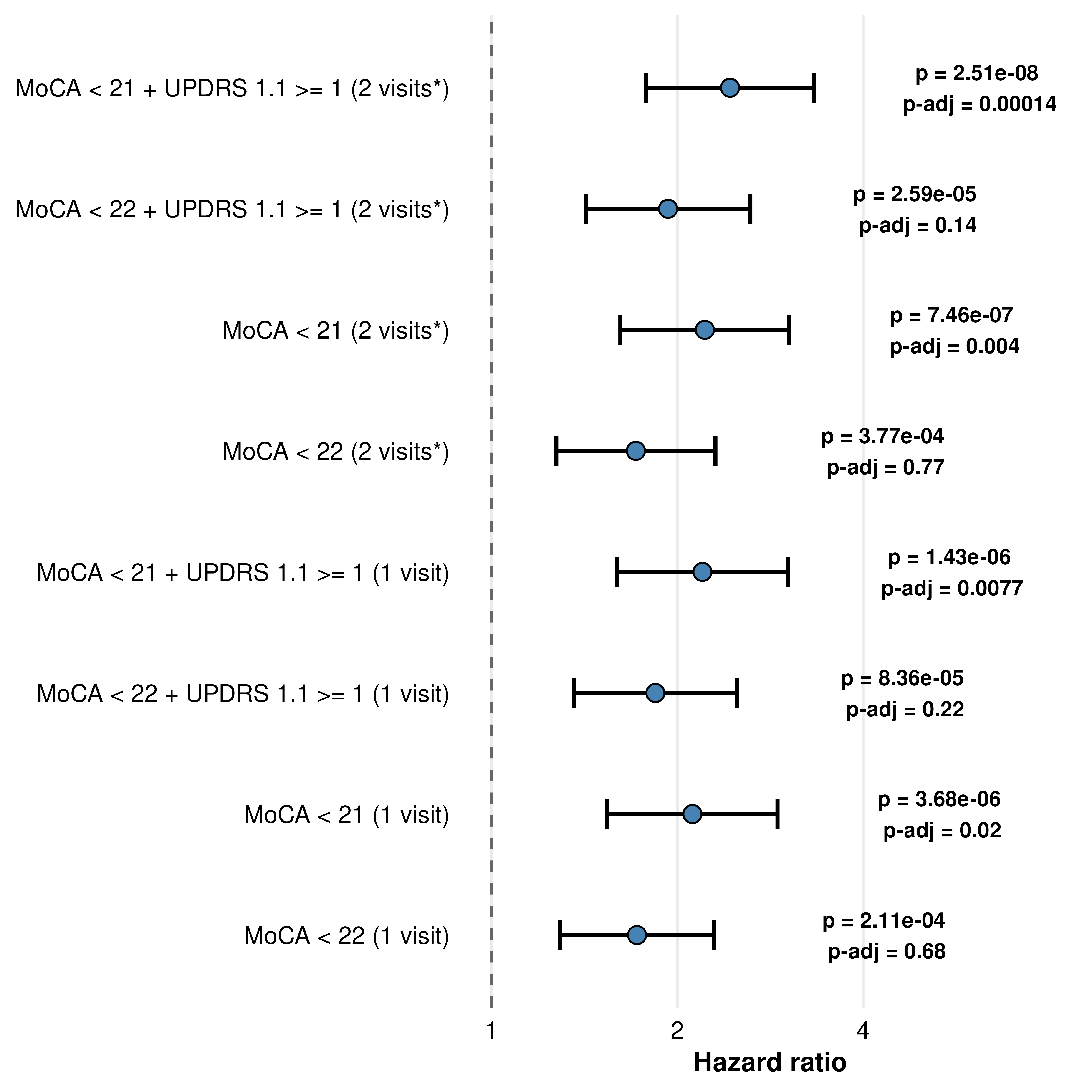

** Met dementia criteria for 2 consecutive where available, otherwise met criteria at last visit*

**Supplementary Figure 8. PPMI Project 293: Sensitivity Analysis**

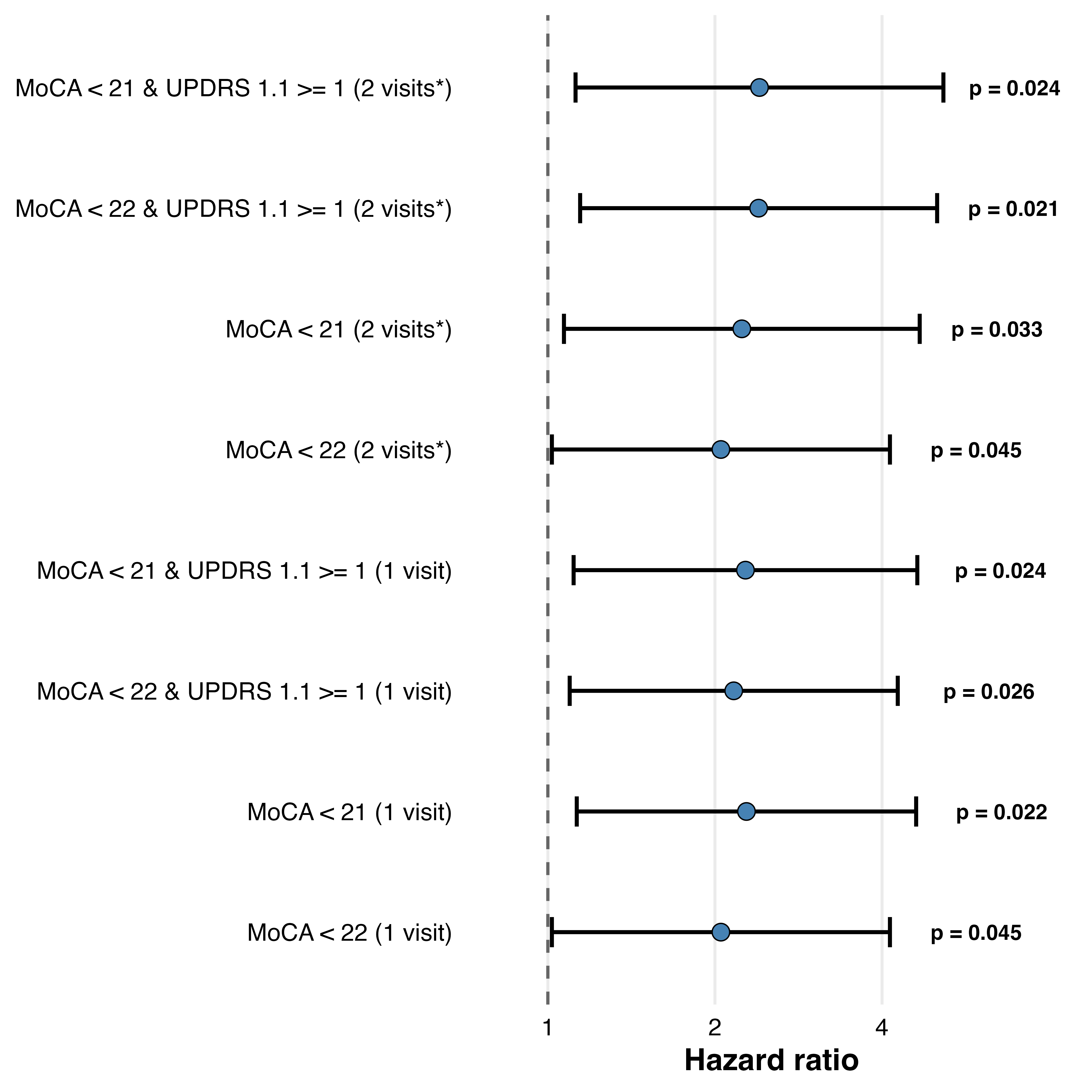

** Met dementia criteria for 2 consecutive where available, otherwise met criteria at last visit*

**Supplementary Figure 9. PPMI Project 277: Sensitivity Analysis**

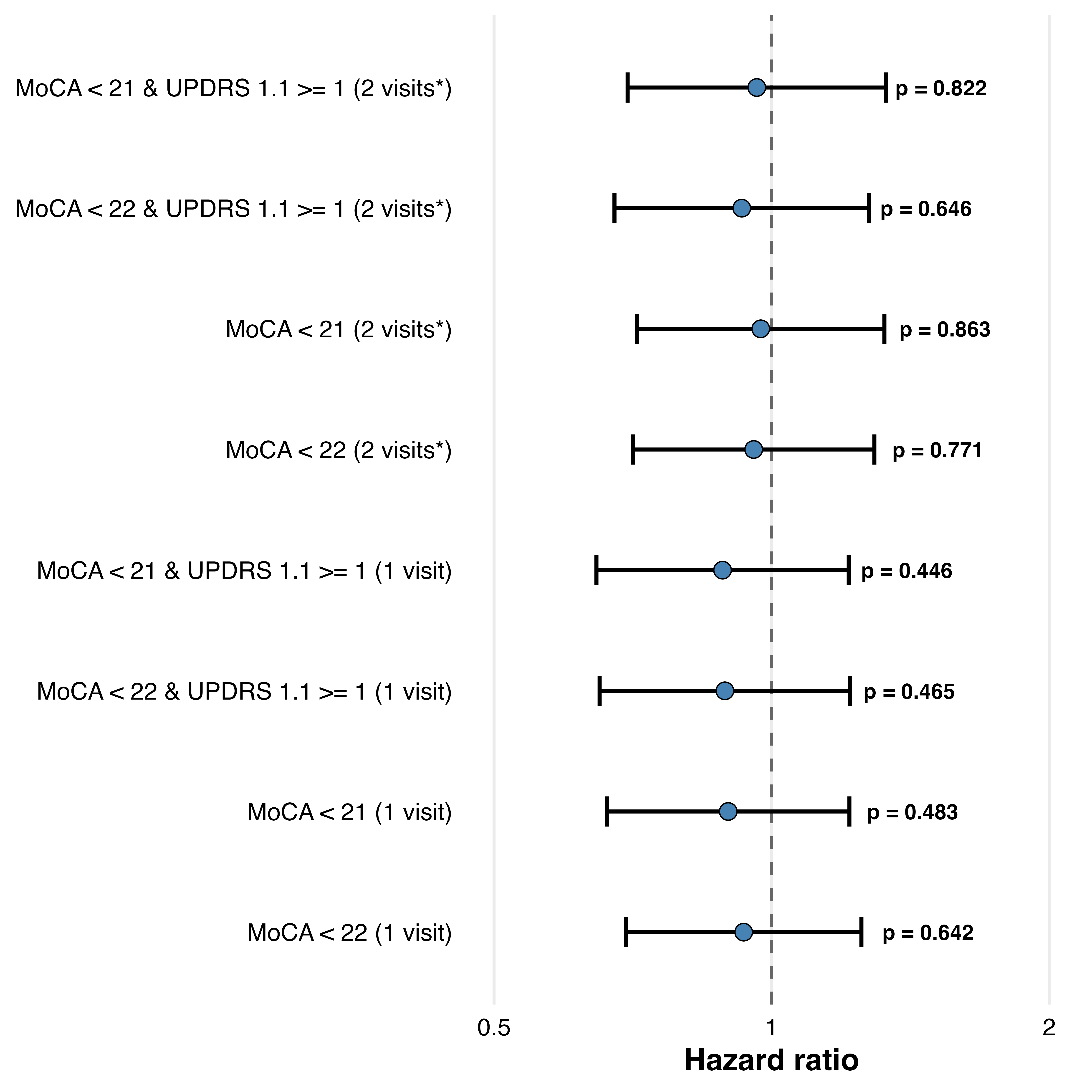

** Met dementia criteria for 2 consecutive where available, otherwise met criteria at last visit*

**Supplementary Figure 10. Correlation between plasma and CSF GFAP in the PPMI cohort**

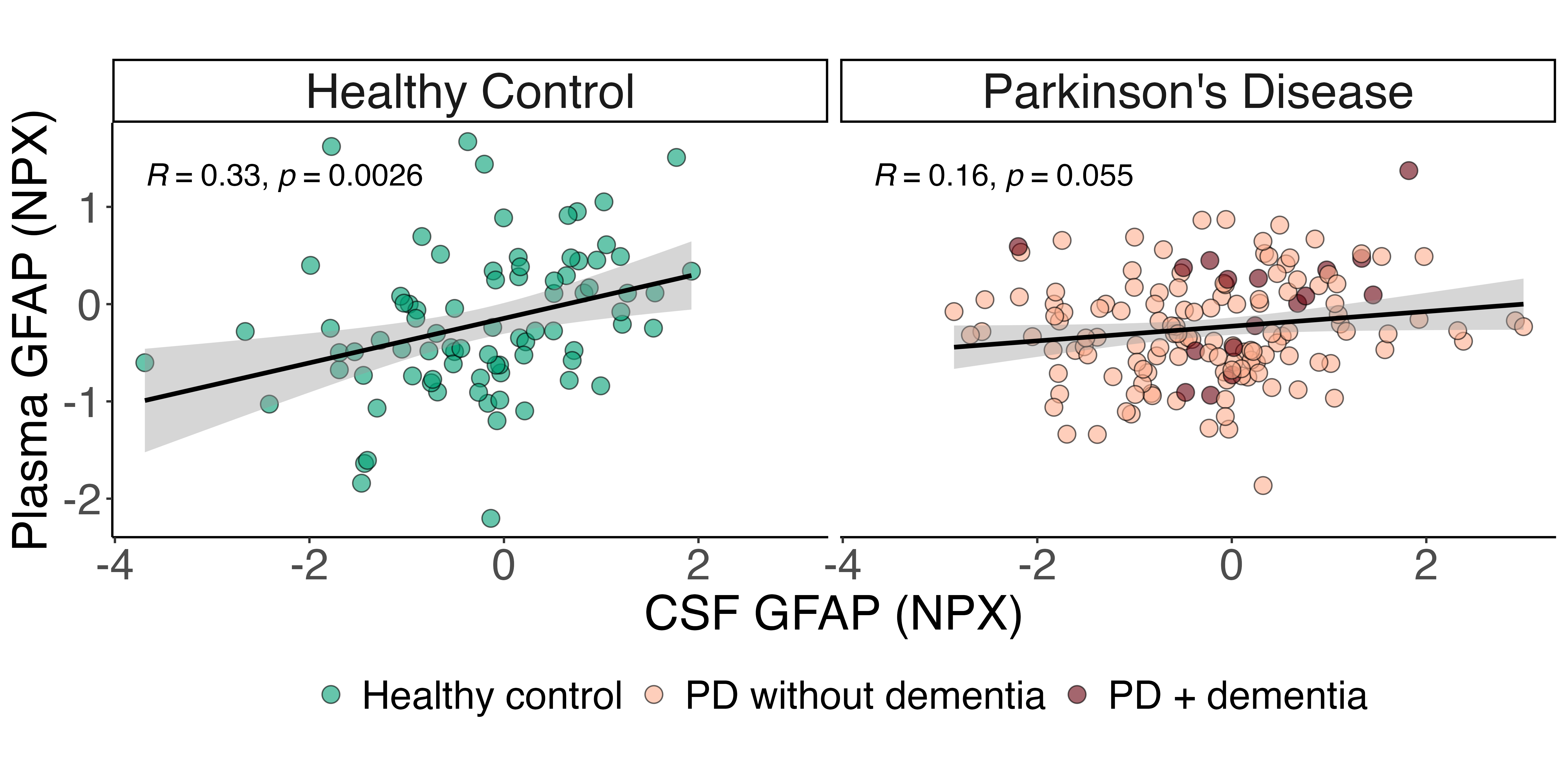

*R = Pearson’s correlation coefficient. P-value derived from a two-sided t-test.*

**Supplementary Figure 11. Associations between GFAP and relevant variables**

**OPDC Discovery cohort**

**(A) GFAP and age	(B) GFAP and sex**

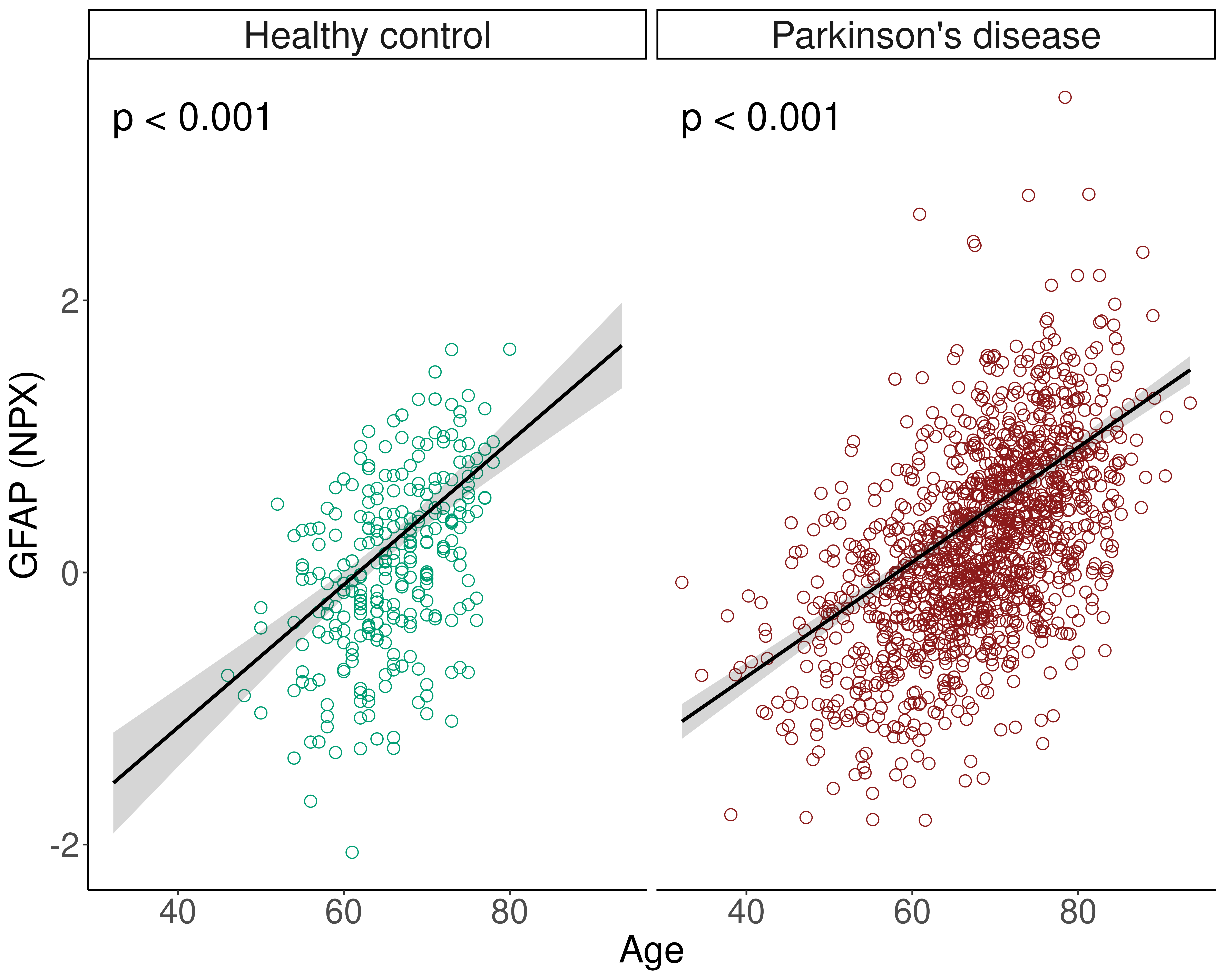

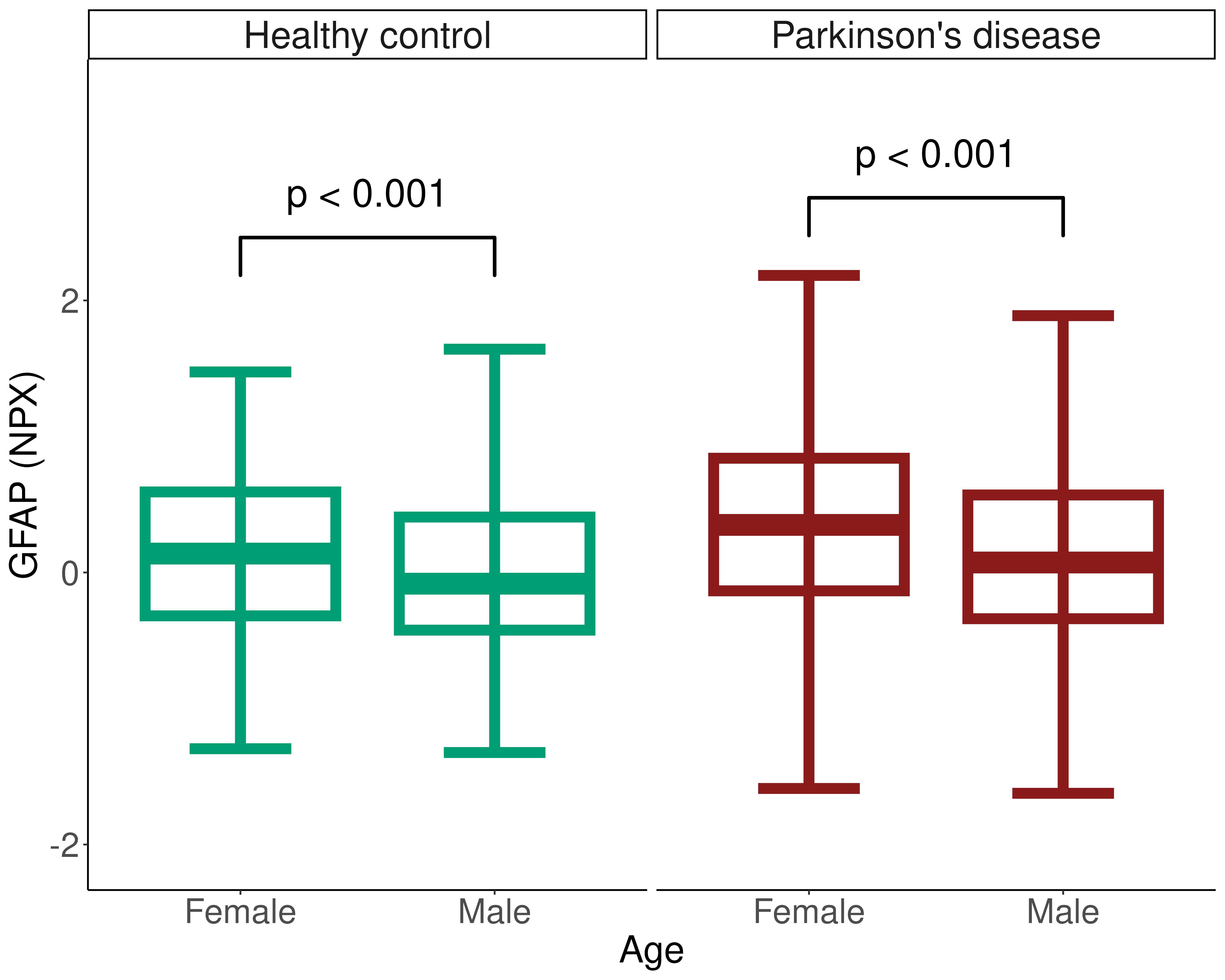

**(C) GFAP and BMI	(D) GFAP and APOE ε4 carrier status**

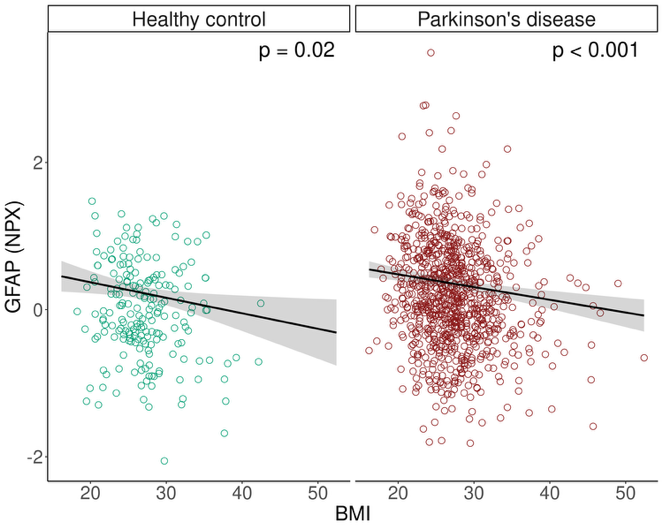

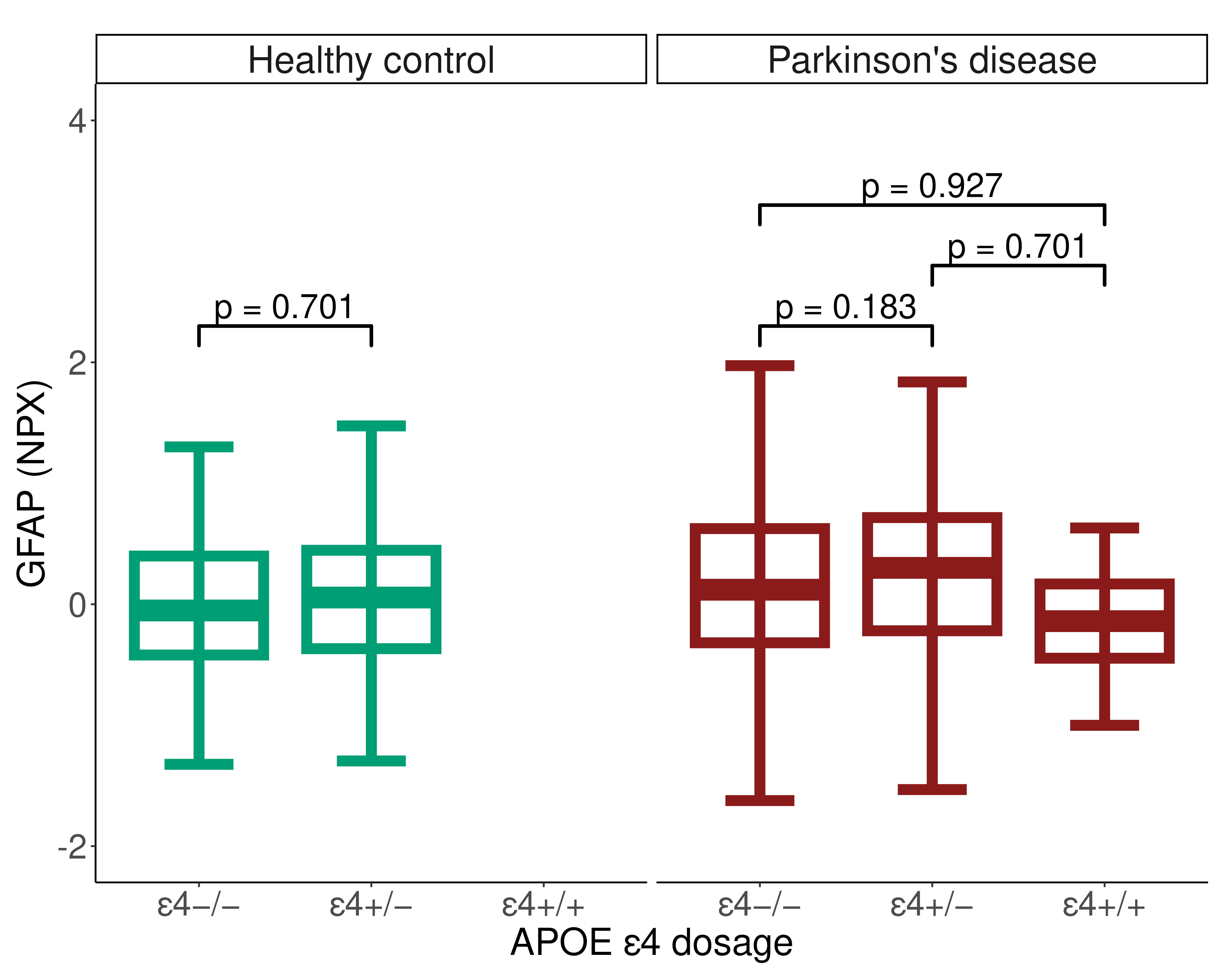

**(E) PD vs HC: OPDC Discovery cohort	(F) PD vs HC: PPMI Project 293**

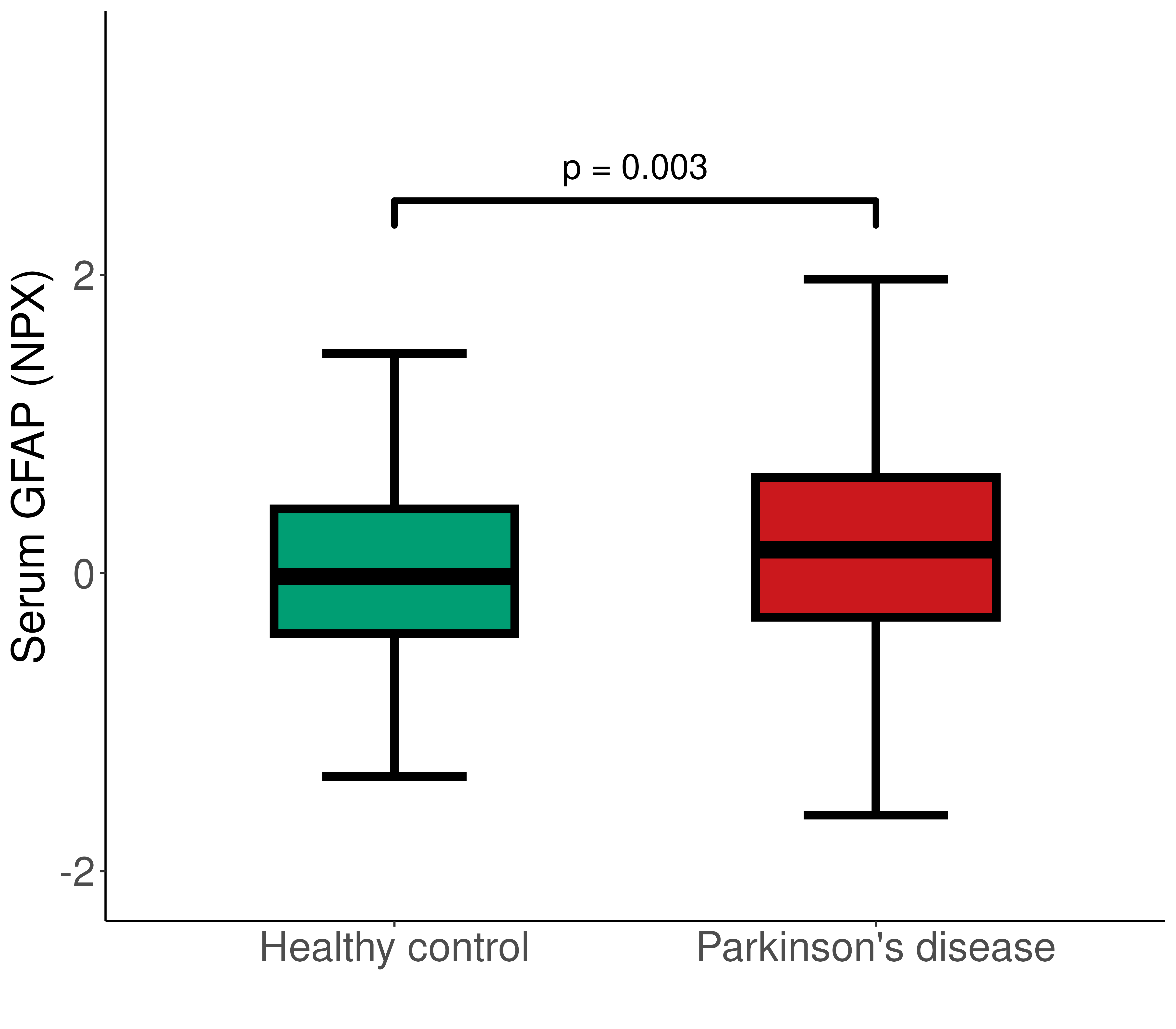

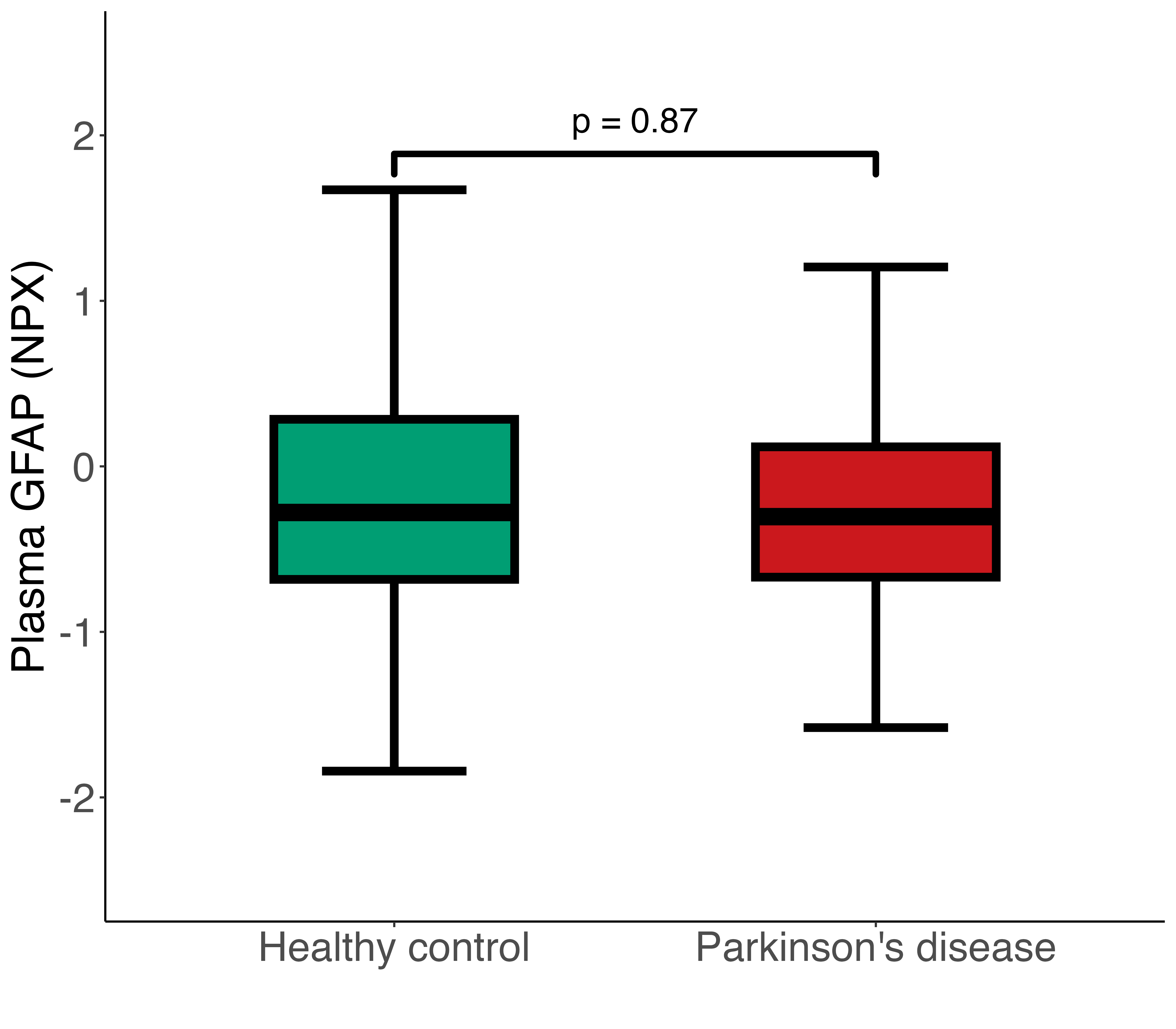

**(G) PD vs HC: PPMI Project 277**

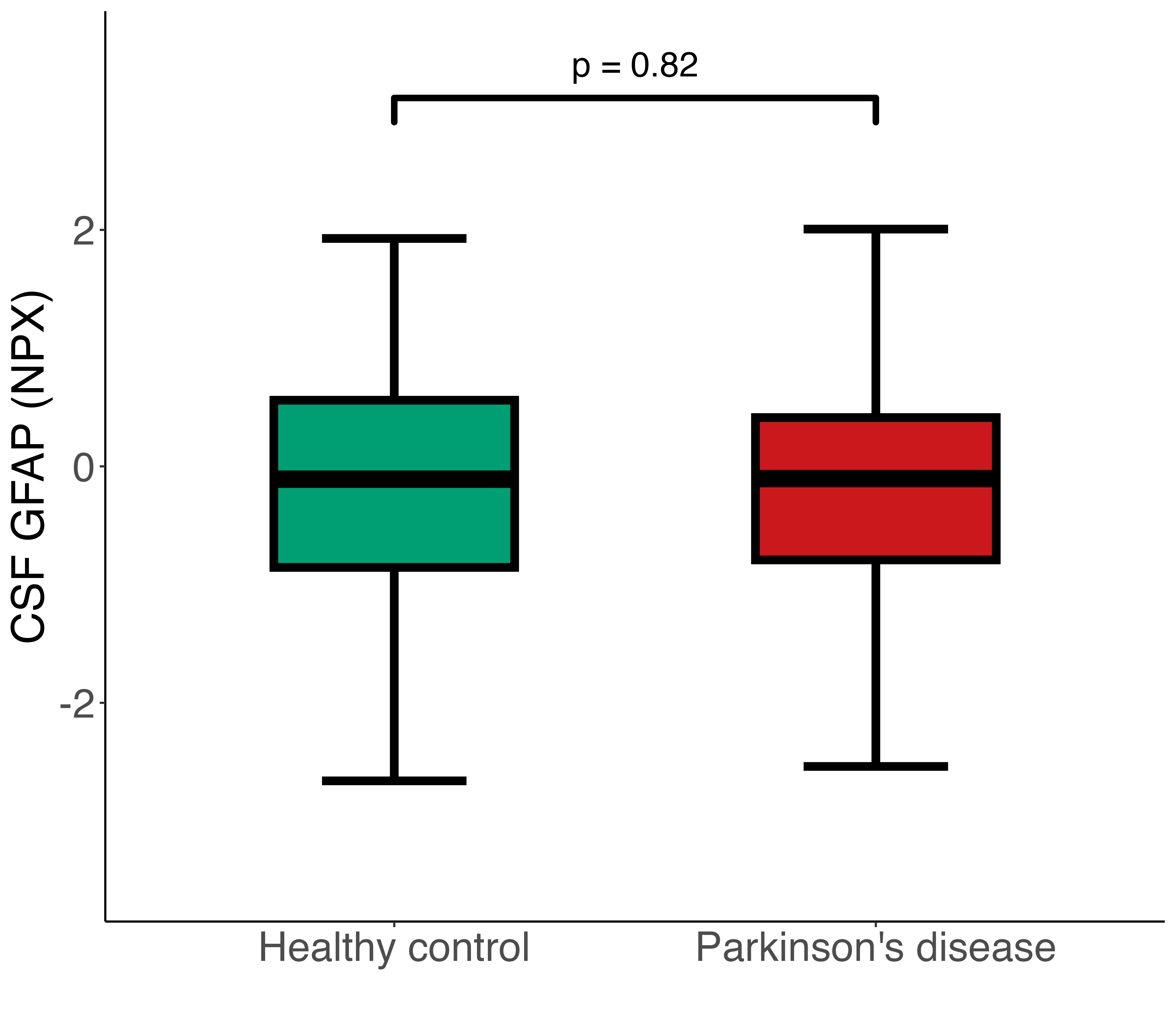

***(A)*** *Raw GFAP NPX displayed. Linear regression of GFAP on age by diagnostic group. Covariates: sex, BMI, APOE ε4 carrier status, sample age.* ***(B)*** *Raw GFAP NPX displayed. Linear regression of GFAP on sex by diagnostic group. Covariates: age, BMI, APOE ε4 carrier status, sample age.* ***(C)*** *Raw GFAP NPX displayed. Linear regression of GFAP on BMI by diagnostic group. Covariates: age, sex, APOE ε4 carrier status, sample age.* ***(D)*** *Raw GFAP NPX displayed. Linear regression of GFAP on APOE ε4 carrier status by diagnostic group. Covariates: age, sex, BMI, sample age.* ***(E)*** *Raw GFAP NPX displayed. Linear regression of GFAP on diagnostic group. Covariates: age, sex, APOE ε4 carrier status , BMI, and sample age.* ***(F-G)*** *Linear regression of GFAP on diagnostic group. Covariates: age, sex and BMI.*
